# Personalized Fluid Management in Patients with Sepsis and AKI: A Casual Machine Learning Approach

**DOI:** 10.1101/2024.08.06.24311556

**Authors:** Wonsuk Oh, Kullaya Takkavatakarn, Zainab Al-taie, Hannah Kittrell, Khaled Shawwa, Hernando Gomez, Ashwin S. Sawant, Pranai Tandon, Gagan Kumar, Michael Sterling, Ira Hofer, Lili Chan, John Oropello, Roopa Kohli-Seth, Alexander W Charney, Monica Kraft, Patricia Kovatch, Mayte Suárez-Fariñas, John A. Kellum, Girish N. Nadkarni, Ankit Sakhuja

## Abstract

**Importance:** Intravenous (IV) fluids are cornerstone for management of acute kidney injury (AKI) after sepsis but can cause fluid overload. Restrictive fluid strategy may benefit some patients, however, identifying them is challenging. Novel causal machine learning (ML) techniques can estimate heterogenous treatments effects (HTE) of IV fluids among these patients.

**Objectives:** To develop and validate causal ML framework to identify patients who benefit from restrictive fluids (<500mL fluids within 24 hours after AKI).

**Design, Setting and Participants:** We conducted a retrospective study among patients with sepsis who developed AKI within 48 hours of ICU admission. We developed a causal ML approach to estimate individualized treatment effects and guide fluid therapy. We developed the model in MIMIC-IV and externally validated in SICdb.

**Main Outcomes and Measures:** Our primary outcome was early AKI reversal at 24 hours. Secondary outcomes included sustained AKI reversal and major adverse kidney events by 30 days (MAKE30). Model performance to identify HTE of restrictive IV fluids was assessed using area under targeting operator characteristic curve (AUTOC), that quantifies how well a model captures HTE and compared to random forest model.

**Results:** Causal forest model outperformed random forest in identifying HTE of restrictive IV fluids with AUTOC 0.15 vs. -0.02 in external validation cohort. Among 1,931 patients in external validation cohort, the model recommended restrictive fluids for 68.9%. Among these, patients who received restrictive fluids demonstrated significantly higher rate of early AKI reversal (53.9% vs 33.2%, p<0.001), sustained AKI reversal (34.2% vs 18.0%, p<0.001) and lower rates of MAKE30 (17.1% vs 34.6%, p=0.003). Results were consistent in adjusted analysis.

**Conclusions and Relevance:** Causal ML framework outperformed random forest model in identifying patients with AKI and sepsis who benefit from restrictive fluid therapy. This provides a data-driven approach for personalized fluid management and merits prospective evaluation in clinical trials.

**KEY POINT.:** *Question:* Can causal machine learning (ML) identify which critically ill, septic patients with AKI benefit from a restrictive fluid management strategy?

*Findings:* In this retrospective cohort study, we developed and externally validated a causal-ML model to identify septic patients with AKI who would benefit from restrictive IV fluid therapy. Those who received the model-recommended restrictive IV fluids experienced significantly higher rates of early AKI reversal (53.9 % vs. 33.2 %; p<0.001) and lower rates of MAKE 30 (17.1% vs 34.6%, p=0.003).

*Meaning:* This study demonstrates that causal ML can effectively identify septic patients with AKI who are most likely to benefit from a restrictive IV fluids strategy.

## INTRODUCTION

Acute kidney injury (AKI) is seen in over one-third of critically ill patients with sepsis and is associated with worse outcomes(1). Oliguria, a frequent clinical indicator of AKI, is common in these patients(2) and is the second most common reason for intravenous (IV) fluids administration(3). Since the primary emphasis is on volume optimization, IV fluids are the cornerstone of AKI management. Nonetheless, this approach might not be optimal in all patients. First, IV fluids do not consistently increase urine output(4). Second, reliance on IV fluid therapy increases the risk of fluid accumulation, frequently resulting in fluid overload. This phenomenon is associated with AKI progression(5–7), and higher mortality(7).

Thus, a more personalized approach to IV fluid administration in critically ill patients is warranted. Restrictive IV fluids in acute lung injury have been associated with shorter duration of mechanical ventilation(8). Similarly, a restrictive fluid approach to resuscitation for septic shock is safe and associated with less fluid overload(9). More recently, restrictive IV fluids in AKI have been associated with lower cumulative fluid balance and reduced dialysis initiation(10). Nevertheless, translating these findings into clinical practice has been challenging due to difficulties in accurately identifying which patients would most benefit from fluid restriction.

Recent advances in causal machine learning (ML) offer a promising avenue to overcome this challenge by enabling heterogeneous treatment effect (HTE) estimation directly from observational data(11, 12). Unlike traditional predictive models, which forecast outcomes under observed conditions, causal ML methods aim to estimate the effect of a specific intervention, such as use of restrictive IV fluids, on an outcome of interest for each patient. This individualized causal inference framework is well-suited to critical care settings, where treatment decisions often need to be tailored to complex and heterogeneous patient populations.

In this study, we used causal ML to estimate HTEs associated with restrictive IV fluid use in critically ill patients with sepsis and AKI, and compared its performance with random forest model. This comparison allows us to evaluate whether causal ML more effectively identifies patients who benefit from restrictive IV fluids strategies. Our goal is to enable a personalized approach to fluid management in this high-risk population.

## METHODS

### Study Design and Data Sources

We conducted a retrospective study using two critical care databases: Medical Information Mart for Intensive Care IV (MIMIC-IV)(13) and Salzburg Intensive Care database (SICdb)(14). MIMIC-IV includes granular data from patients in intensive care units (ICU) at Beth Israel Deaconess Medical Center from 2008-2022, and SICdb contains ICU data from University Hospital Salzburg, Austria from 2013-2021. MIMIC-IV served as the development cohort, and SICdb was used for external validation.

### Study Population

We included adults aged 18 years or older, who developed sepsis within 24 hours of admission to intensive care unit (ICU)(15), and developed AKI within 48 hours after admission to ICU. We identified sepsis in accordance with third international consensus definition(16) as a change of SOFA score of 2 or more, in patients with suspected infection. Suspected infection was defined as a combination of blood cultures and antibiotic administration such that if blood culture was performed first, the antibiotic must be given within 72 hours and if antibiotic was given first then blood culture should be done within 24 hours(16). In accordance with Kidney Disease Improving Global Outcomes (KDIGO) guidelines(17), we defined AKI as either a rise in serum creatinine of 0.3 mg/dL or more within 48 hours or an increase of at least 1.5 times the reference serum creatinine within 7 days, or a urine output of less than 0.5 ml/kg/h for at least a 6-hour time-period. We have provided further details for identification of AKI in **Supplementary Methods**. We considered only the first ICU admission for those patients with multiple admissions. We excluded patients with a history of end-stage kidney disease or kidney transplant. We also excluded patients without serum creatinine measurements within 24 hours of AKI onset. Patients who died or were transferred out of the ICU within 24 hours of AKI onset were excluded, as they would not have had sufficient opportunity to receive the intervention. (**Supplementary Fig S1**).

### Outcomes

The primary outcome of our study was early AKI reversal defined as the patient no longer meeting KDIGO criteria for AKI at 24 hours of AKI onset(17). The secondary outcomes included sustained AKI reversal, defined as maintaining AKI reversal for 48 hours or longer(18), and major adverse kidney events by 30 days (MAKE30) defined as a composite of in-hospital death, new dialysis or persistent kidney dysfunction within 30 days of ICU admission or by hospital discharge, whichever occurred first (19). In consistence with prior literature, persistent kidney dysfunction was defined as the final inpatient serum creatinine value as ≥200% of reference creatinine(19).

### Treatment

The treatment of interest in this study was the amount of IV fluids administered, including both crystalloids and colloids, from the onset of AKI through the first 24 hours afterward. We defined ‘restrictive fluid strategy’ as administrating less than 500mL of IV fluids within this specified time frame.

### Features

We extracted data on patient demographics, such as age, sex, and race/ethnicity. We also extracted data for vital signs, SOFA scores, vasopressor administration, use of mechanical ventilation, administration of nephrotoxic medications(20), urine output and net fluid balance from ICU admission to onset of AKI. Net fluid balance was calculated as the difference between all fluid inputs (including crystalloids, colloids, medications, transfusions, and flushes) and outputs (such as urine, drain outputs, and other measured losses). Laboratory values were extracted from ICU admission to AKI onset, as well as from the 48-hour period prior to AKI onset, including values obtained before ICU admission. Additionally, we collected data on treatment - the amount of IV fluids administered from the onset of AKI through the first 24 hours afterward. Physiologically improbable values were removed(21). For vital signs, laboratory values and SOFA scores, we collected the highest, lowest and latest values when multiple values were present. We only included features that were present in over 70% of the cohort. Missing data were imputed using the multivariate imputation by chained equations (MICE) method(22).

### Development of Causal ML Approach

We used causal ML, a novel machine learning strategy to identify critically ill septic patients with AKI who would benefit from a restrictive fluid strategy. This employed a two-step approach. First, we applied causal forests, a non-parametric extension of random forests, to estimate HTEs across patients(23). Causal forests provide individualized treatment effect (ITE) estimates while adjusting for confounding through sample splitting and inverse probability weighting while using a doubly robust approach(24). These estimates help identify variations in responses to IV fluid restriction (the HTEs), that are not captured by average treatment effects alone. Subsequently, we applied policy tree algorithm to translate these individualized effects into an interpretable set of clinical decision rules(25). Policy trees use recursive partitioning to stratify patients based on shared clinical features, producing simple, actionable guidelines that can inform bedside decisions. We have provided details of causal forest and policy tree methodology in **Supplementary Methods**.

### Statistical Analysis

We expressed continuous features as mean and standard deviation and categorical features as proportions. We used ANOVA or chi-square tests for comparisons as appropriate. We used logistic regression models to evaluate associations between restrictive fluid therapy and clinical outcomes. To evaluate how effectively our causal forest model captured HTEs, we computed the area under the targeting operator characteristic curve (AUTOC) (26). AUTOC quantifies the observed benefit of treatment, restrictive IV fluid administration, on the outcome of early AKI reversal, when patients are ranked according to the magnitude of their estimated ITEs. AUTOC represents the model’s ability to capture the heterogeneous effects of restrictive IV fluids on the risk of early AKI reversal for individual patients. Thus, a higher AUTOC indicates a stronger ability to identify patients who benefit the most from treatment. To evaluate the added value of causal ML in identification of HTEs for restrictive IV fluids, we compared the AUTOC of causal forest model to that of a conventional random forest model(27) trained to predict the outcome. This comparison enabled us to evaluate whether the causal model more effectively captured meaningful HTE. To ensure robustness of our findings, we also performed sensitivity analyses. Detailed descriptions of statistical methodology and sensitivity analyses are provided in **Supplementary Methods**.

All analyses were conducted using R (v4.3.0). We followed the TRIPOD (Transparent Reporting of a multivariable prediction model for Individual Prognosis Or Diagnosis) AI guideline(28) for reporting clinical prediction models study (**Supplementary Table S1**). The study received approval from the Institutional Review Board at the Icahn School of Medicine at Mount Sinai (approval no. 19-00951).

## RESULTS

### Study Population

Our study included 11,650 patients from the development cohort (MIMIC-IV), and 1,931 patients from the external validation cohort (SICdb). The average age in the development cohort was 67.1 ± 15.8 years with 58.0% males. The average age in the external validation cohort was 66.9 ± 15.3 years with 62.2% males. The baseline characteristics are shown in **Table 1** and **Table S2** in the online data supplement.

**Table 1.** Comparison of baseline characteristics of patients identified by the policy tree as likely to benefit versus not benefit from restrictive fluids.

|  | Development cohort (MIMIC-IV) |  |  | External validation cohort (SICdb) |  |  |
| --- | --- | --- | --- | --- | --- | --- |
|  | Identified to have no benefit from restrictive fluid strategy (N= 6,526) | Identified to benefit from restrictive fluid strategy (N= 5,124) | p value | Identified to have no benefit from restrictive fluid strategy (N= 600) | Identified to benefit from restrictive fluid strategy (N= 1,331) | p value |
| Age, year, (mean±SD) | 66.4 ± 15.9 | 68.0 ± 15.7 | < 0.001 | 66.7 ± 15.6 | 67.1 ± 15.2 | 0.6 |
| Gender - Male, n (%) | 3860 (59.1%) | 2892 (56.4%) | 0.003 | 387 (64.5%) | 814 (61.2%) | 0.2 |
| Race - White, n(%) | 4274 (65.5%) | 3377 (65.9%) | 0.7 | NA<sup>C</sup> | NA<sup>C</sup> |  |
| Race - Black, n(%) | 597 (9.1%) | 472 (9.2%) |  | NA<sup>C</sup> | NA<sup>C</sup> |  |
| Race - Hispanic, n(%) | 193 (3.0%) | 166 (3.2%) |  | NA<sup>C</sup> | NA<sup>C</sup> |  |
| Race - Other, n(%) | 1462 (22.4%) | 1109 (21.6%) |  | NA<sup>C</sup> | NA<sup>C</sup> |  |
| Congestive heart failure, n (%) | 4211 (64.5%) | 3266 (63.7%) | 0.4 | NA<sup>C</sup> | NA<sup>C</sup> |  |
| Chronic kidney disease, n (%) | 1355 (20.8%) | 1009 (19.7%) | 0.2 | 100 (16.7%) | 203 (15.3%) | 0.5 |
| Baseline creatinine, mg/dL, (mean±SD) | 1.1 ± 0.6 | 1.1 ± 0.6 | 0.8 | 1.0 ± 0.5 | 1.0 ± 0.5 | 0.2 |
| AKI stage - 1, n (%) | 5629 (86.3%) | 4491 (87.6%) | 0.05 | 536 (89.3%) | 1225 (92.0%) | 0.05 |
| AKI stage - 2, n (%) | 532 (8.2%) | 358 (7.0%) |  | 20 (3.3%) | 45 (3.4%) |  |
| AKI stage - 3, n (%) | 365 (5.6%) | 275 (5.4%) |  | 44 (7.3%) | 61 (4.6%) |  |
| SOFA at the time of sepsis onset, (mean±SD) | 4.2 ± 2.3 | 4.5 ± 2.4 | < 0.001 | 4.02 ± 1.8 | 4.2 ± 2.0 | 0.05 |
| Vasopressors<sup>B</sup>, n (%) | 2542 (39.0%) | 2391 (46.7%) | < 0.001 | 386 (64.3%) | 1049 (78.8%) | < 0.001 |
| Vasopressors<sup>B</sup>, hours (mean±SD) | 3.3 ± 6.4 | 4.2 ± 7.2 | < 0.001 | 4.5 ± 5.8 | 7.0 ± 7.8 | < 0.001 |
| Mechanical ventilation<sup>B</sup>, n (%) | 3671 (56.3%) | 2983 (58.2%) | 0.03 | 289 (48.2%) | 619 (46.5%) | 0.5 |
| Mechanical ventilation<sup>B</sup>, hours (mean±SD) | 6.4 ± 8.6 | 6.5 ± 8.5 | 0.2 | 4.0 ± 5.9 | 4.6 ± 7.3 | 0.08 |
| Nephrotoxic drugs administration<sup>A</sup>, n (%) | 1833 (28.1%) | 1433 (28.0%) | 0.9 | 190 (31.7%) | 481 (36.1%) | 0.06 |
| ICU length of stay, (mean±SD) | 6.9 ± 7.6 | 7.2 ± 7.3 | 0.02 | 7.8 ± 9.0 | 8.7 ± 10.5 | 0.09 |
| Hospital length of stay, (mean±SD) | 14.5 ± 13.9 | 14.5 ± 13.0 | 0.9 | 24.5 ± 21.6 | 25.7 ± 21.2 | 0.2 |
| Inhospital mortality, n (%) | 1243 (19.0%) | 1063 (20.7%) | 0.02 | 185 (30.8%) | 423 (31.8%) | 0.7 |

Abbreviation: AKI (acute kidney injury), SOFA (sequential organ failure assessment)
<sup>A</sup>: Measurements from the last 48 hours prior to AKI onset
<sup>B</sup>: Measurements from ICU admission to AKI onset
<sup>C</sup>: NA – Not Available

### Robustness of HTE estimation by Causal Forest

To assess the performance of causal forest for estimation of HTE, we compared the AUTOC for both causal forest and conventional random forest models (**Fig 1A and 1B**).

**Figure 1.**
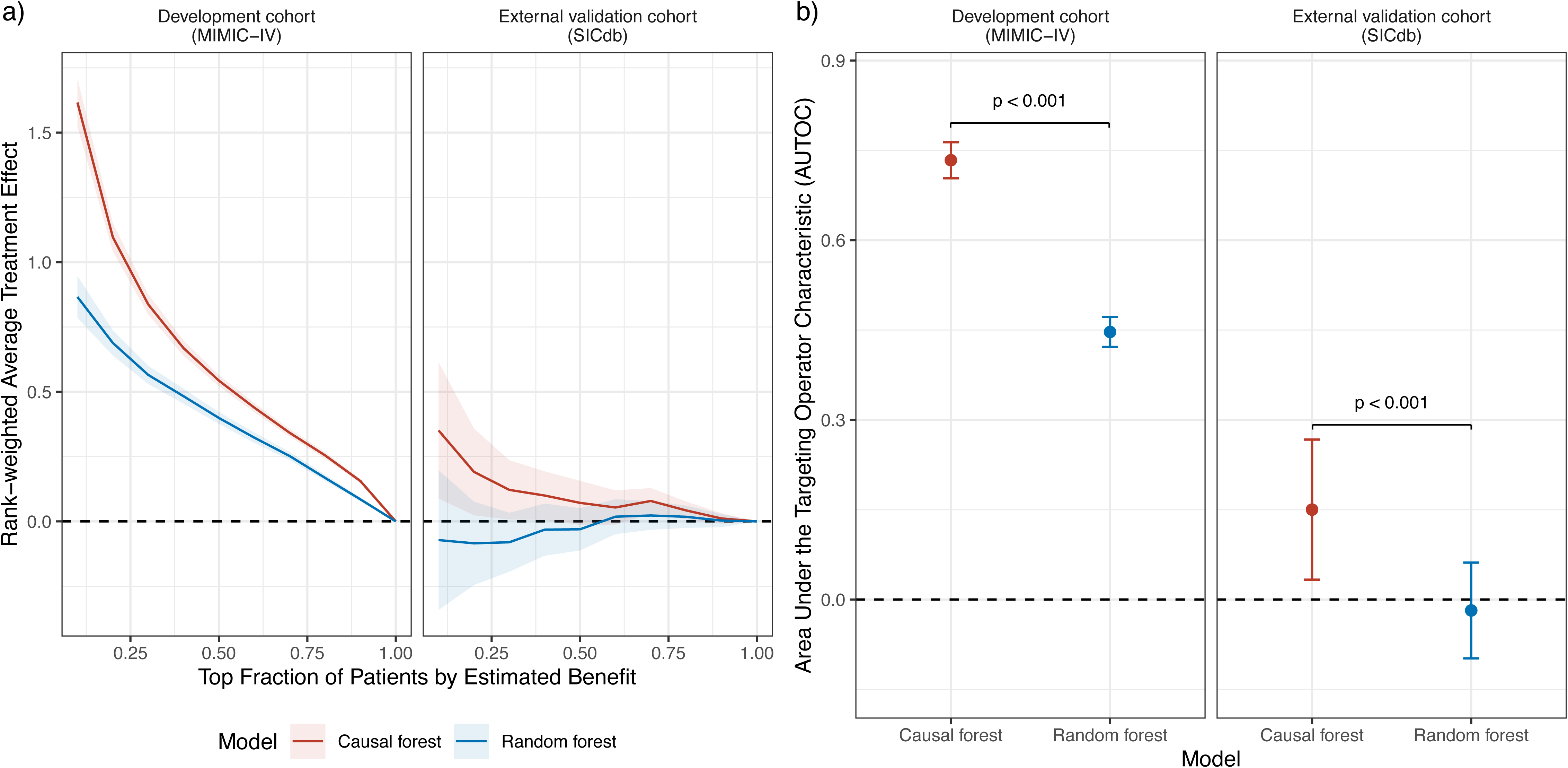
Comparison of causal and random forest models: a) AUTOC scores for individual treatment effects in development and validation cohorts (p-values from two-sample t-test); b) Quantile-based distributions of estimated treatment effects by causal forest and random forest models.

In the development cohort, the causal forest model demonstrated an AUTOC of 0.73 (95% CI: 0.70 – 0.76), substantially outperforming the noncausal random forest model, which achieved an AUTOC of 0.45 (95% CI: 0.42 – 0.47). In the external validation cohort, we observed consistent findings. The causal forest model achieved an AUTOC of 0.15 (95% CI: 0.03 – 0.27), compared to an AUTOC of -0.02 (95% CI: -0.10 – 0.06) for the noncausal random forest. Overall causal models captured HTE better and were validated in external cohort, while RF did not.

### Policy Tree for Restrictive Fluid Strategy in Critically Ill Septic Patients with AKI

Our policy tree uses ITEs estimated by causal forest model to identify patients with AKI and sepsis who are likely to benefit from a restrictive fluid strategy(**Fig. 2**). The key determinants identified for early AKI reversal were respiratory rate, systolic blood pressure (SBP), weight, platelet, temperature, blood urea nitrogen (BUN), partial pressure of carbon dioxide (pCO2), mean corpuscular hemoglobin (MCH), diastolic blood pressure (DBP), partial pressure of oxygen (pO2), heart rate, red blood cell (RBC) count, mean corpuscular hemoglobin concentration (MCHC), age, hematocrit, and urine output.

**Figure 2.**
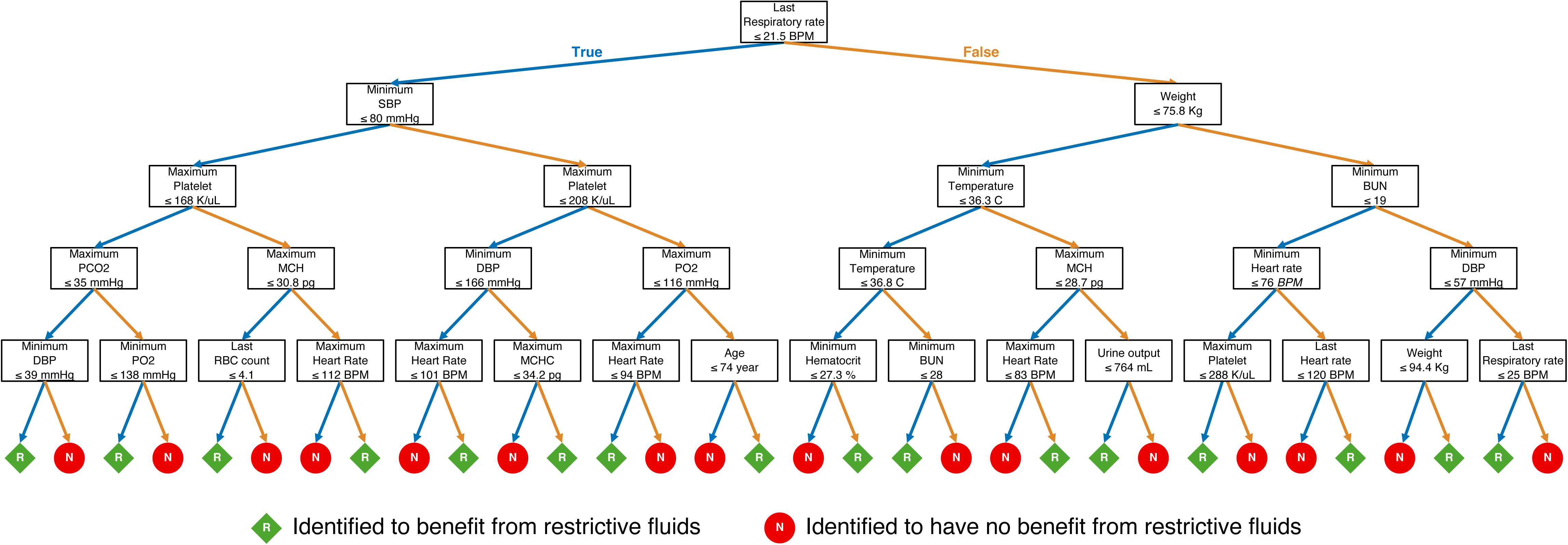
Policy Tree for Restrictive Fluid Strategy in Septic Patients with Acute Kidney Injury. Legend: Features: Age, Blood Urea Nitrogen (BUN), Diastolic Blood Pressure (DBP), Heart rate, Hematocrit, Mean Corpuscular Hemoglobin (MCH), Mean Corpuscular Hemoglobin Concentration (MCHC), Partial Pressure of Carbon Dioxide (PCO2), Partial Pressure of Oxygen (PO2), Red Blood Cell (RBC) count, Platelet, Respiratory rate, Systolic Blood Pressure (SBP), Weight, Temperature, Urine output

The policy tree suggested benefit from a restrictive fluid strategy after development of AKI among 5,124 (44.0%) patients in the development cohort and 1,331 (68.9%) patients in the external validation cohort. In development cohort, patients identified to benefit from restrictive fluid strategy were more often receiving vasopressors (46.7% vs. 39.0%, p<0.001) and mechanical ventilation (58.2% vs. 56.3%, p=0.03). In external validation cohort, these patients were also more frequently on vasopressors (78.8% vs. 64.3%, p<0.001) but there was no statistically significant difference among those on mechanical ventilation (46.5% vs. 48.2%, p=0.5). The baseline characteristics of these patients are shown in **Table 1** and **Supplementary Table S2**. Among the patients identified to benefit from restrictive fluid therapy, only 798 (15.6%) patients in the development cohort and 76 (5.7%) patients in the external validation cohort received restrictive fluid therapy. In the development cohort, among patients identified as likely to benefit from a restrictive fluid strategy, those who actually received restrictive fluids were less frequently on vasopressors (15.4% vs. 52.4%; p<0.001) and mechanical ventilation (43.1% vs. 61.0%; p<0.001). In contrast, in the external validation cohort, there were no statistically significant differences in vasopressor use (71.1% vs. 79.3%; p=0.09) or mechanical ventilation (51.3% vs. 46.2%; p=0.40) among these patients(**Table 2 and Table S3** in the online data supplement).

**Table 2.** Comparison of additional baseline characteristics of patients who actually received versus did not receive restrictive fluids within the subgroup predicted to benefit from restrictive fluids.

|  | Development cohort (MIMIC-IV) |  |  | External validation cohort (SICdb) |  |  |
| --- | --- | --- | --- | --- | --- | --- |
|  | Did not receive restrictive IV fluids<br>(N= 4,326) | Received restrictive IV fluids<br>(N= 798) | p value | Did not receive restrictive IV fluids<br>(N= 1255) | Received restrictive IV fluids<br>(N= 76) | p value |
| Age, year, (mean±SD) | 67.7 ± 15.7 | 69.7 ± 15.1 | < 0.001 | 67.1 ± 15.2 | 67.1 ± 14.2 | 1.0 |
| Gender - Male, n (%) | 2458 (56.8%) | 434 (54.4%) | 0.2 | 767 (61.1%) | 47 (61.8%) | 0.9 |
| Race - White, n(%) | 2883 (66.6%) | 494 (61.9%) | 0.02 | NA<sup>C</sup> | NA<sup>C</sup> |  |
| Race - Black, n(%) | 378 (8.7%) | 94 (11.8%) |  | NA<sup>C</sup> | NA<sup>C</sup> |  |
| Race - Hispanic, n(%) | 139 (3.2%) | 27 (3.4%) |  | NA<sup>C</sup> | NA<sup>C</sup> |  |
| Race - Other, n(%) | 926 (21.4%) | 183 (22.9%) |  | NA<sup>C</sup> | NA<sup>C</sup> |  |
| Congestive heart failure, n (%) | 2598 (60.1%) | 668 (83.7%) | < 0.001 | NA<sup>C</sup> | NA<sup>C</sup> |  |
| Chronic kidney disease, n (%) | 807 (18.7%) | 202 (25.3%) | < 0.001 | 189 (15.1%) | 14 (18.4%) | 0.5 |
| Baseline creatinine, mg/dL, (mean±SD) | 1.1 ± 0.6 | 1.2 ± 0.7 | < 0.001 | 1.0 ± 0.5 | 1.0 ± 0.4 | 0.8 |
| AKI stage - 1, n (%) | 3808 (88.0%) | 683 (85.6%) | 0.1 | 1153 (91.9%) | 72 (94.7%) | 0.6 |
| AKI stage - 2, n (%) | 295 (6.8%) | 63 (7.9%) |  | 43 (3.4%) | 2 (2.6%) |  |
| AKI stage - 3, n (%) | 223 (5.2%) | 52 (6.5%) |  | 59 (4.7%) | 2 (2.6%) |  |
| SOFA at the time of sepsis onset, (mean±SD) | 4.6 ± 2.5 | 3.8 ± 2.0 | < 0.001 | 4.2 ± 2.0 | 3.8 ± 1.8 | 0.04 |
| Vasopressors<sup>B</sup>, n (%) | 2268 (52.4%) | 123 (15.4%) | < 0.001 | 995 (79.3%) | 54 (71.1%) | 0.09 |
| Vasopressors<sup>B</sup>, hours (mean±SD) | 4.6 ± 7.4 | 1.6 ± 5.3 | < 0.001 | 7.0 ± 7.7 | 6.5 ± 9.6 | 0.6 |
| Mechanical ventilation<sup>B</sup>, n (%) | 2639 (61.0%) | 344 (43.1%) | < 0.001 | 580 (46.2%) | 39 (51.3%) | 0.4 |
| Mechanical ventilation<sup>B</sup>, hours (mean±SD) | 6.8 ± 8.6 | 4.8 ± 7.7 | < 0.001 | 4.5 ± 7.1 | 6.2 ± 9.9 | 0.04 |
| Nephrotoxic drugs administration<sup>A</sup>, n (%) | 1117 (25.8%) | 316 (39.6%) | < 0.001 | 453 (36.1%) | 28 (36.8%) | 0.9 |
| ICU length of stay, (mean±SD) | 7.5 ± 7.4 | 5.8 ± 6.4 | < 0.001 | 8.9 ± 10.7 | 5.1 ± 6.1 | 0.003 |
| Hospital length of stay, (mean±SD) | 14.6 ± 12.7 | 13.7 ± 14.4 | 0.07 | 26.0 ± 21.4 | 21.0 ± 15.4 | 0.05 |
| Inhospital mortality, n (%) | 936 (21.6%) | 127 (15.9%) | < 0.001 | 412 (32.8%) | 11 (14.5%) | < 0.001 |

Abbreviation: AKI (acute kidney injury), SOFA (sequential organ failure assessment),
<sup>A</sup>: Measurements from the last 48 hours prior to AKI onset
<sup>B</sup>: Measurements from ICU admission to AKI onset
<sup>C</sup>: NA – Not Available

### Outcomes

In both development and external validation cohorts, patients suggested by policy tree to benefit from a restrictive fluid strategy who received restrictive fluids had significantly higher rates of early AKI reversal in comparison to those that were suggested to have a benefit from restrictive fluids but did not receive them (58.9% vs. 35.8%; p<0.001 in development cohort; 53.9% vs. 33.2%; p<0.001 in external validation cohort) (**Fig. 3a**). On unadjusted analysis, restrictive fluid strategy among patients identified to benefit from restrictive fluids, was associated with significantly greater odds of early AKI reversal in both development (OR 2.57; 95% CI: 2.20 – 3.00) and external validation (OR 2.35; 95% CI: 1.48 – 3.75) cohorts (**Fig. 3b**). This effect was consistent in analysis adjusted for age, sex, race, reference serum creatinine, SOFA score and AKI stage at the time of AKI onset, and net fluid balance from ICU admission to onset of AKI, where restrictive fluid strategy among patients suggested to benefit from restrictive fluids was associated with greater odds of early AKI reversal in both development (OR 2.75; 95% CI: 2.32 – 3.25) and external validation cohorts (OR 2.22; 95% CI: 1.37 – 3.60) (**Fig. 3b**). In comparison, in both development and external validation cohorts, patients identified to have no benefit from a restrictive fluid strategy did not show any improvement in outcomes when they received restrictive fluids (**Fig. 3a and 3b**).

**Figure 3.**
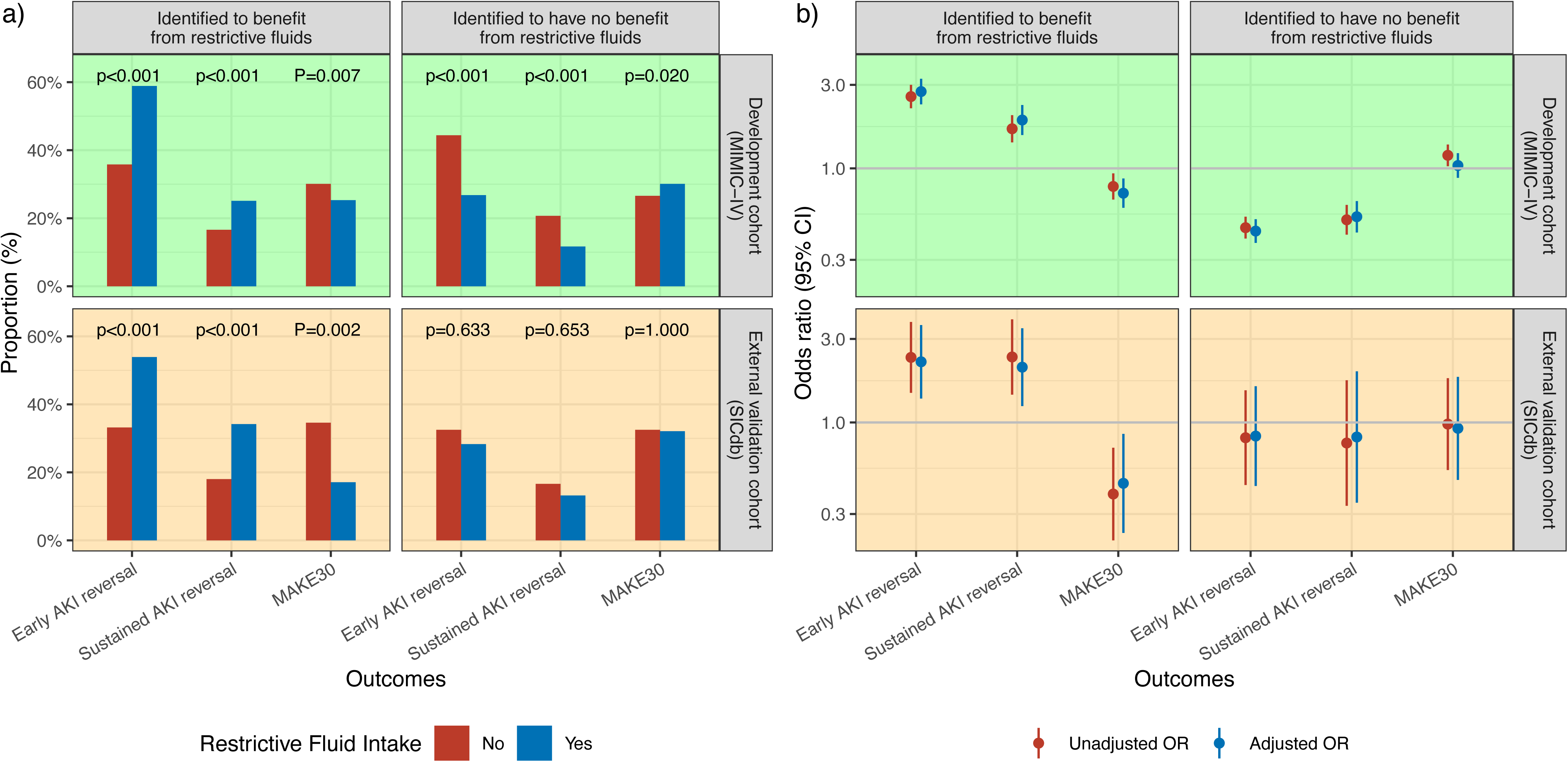
Impact of restrictive fluid strategy among patients stratified by predicted benefit from receipt of restrictive fluid strategy: a) Proportion (p-values from chi-square tests), b) Odds ratio.

Similarly, among patients identified by our causal ML framework as likely to benefit from a restrictive fluid strategy, those who actually received restrictive fluids had higher rates of sustained AKI reversal and lower rates of MAKE30 compared to those who did not, in both the development and external validation cohorts (**Fig. 3a**). In the external validation cohort, among patients identified to benefit from restrictive fluids, 34.2% of those who received restrictive fluids achieved sustained AKI reversal, compared to only 18.0% of those who did not receive them (p<0.001). Additionally, only 17.1% of patients who received restrictive fluids developed MAKE30, compared to 34.6% of those who did not receive them (p=0.003). In the unadjusted analysis of the external validation cohort, use of restrictive fluids among patients identified to benefit from a restrictive fluid strategy was associated with a significantly higher odds sustained AKI reversal (OR 2.37; 95% CI: 1.44 – 3.89) and lower odds of developing MAKE30 (OR 0.39; 95% CI: 0.21 – 0.72). This effect remained consistent in the adjusted analysis, where a restrictive fluid strategy among patients identified to benefit from it was associated with significantly higher odds of sustained AKI reversal (OR 2.; 95% CI: 1.24 – 3.45) and lower odds of MAKE30 (OR 0.45; 95% CI: 0.23 – 0.86). Patients identified as not deriving any benefit from restrictive fluid strategy again showed no benefit from receiving restrictive fluids in both adjusted and unadjusted analyses (**Fig. 3a and 3b**).

Sensitivity analyses demonstrated consistent findings across patients with chronic kidney disease, those with congestive heart failure, and when baseline creatinine was redefined as the admission value for patients without pre-admission measurements. Full details of these analyses are provided in Supplementary Materials.

## DISCUSSION

In this study we have developed and validated a novel, data-driven, causal ML strategy to identify critically ill patients with sepsis and AKI who would benefit from a restrictive fluid strategy. Using large, heterogenous datasets, we demonstrated that patients predicted to benefit from restrictive fluid administration experienced significantly higher rates of early and sustained AKI reversal, and lower rates of MAKE30, when they received fluid-restrictive care. These findings remained robust after adjustment for confounders and were consistent across both internal and external validation cohorts.

Administration of IV fluids to improve cardiac output and consequently kidney blood flow, has been a cornerstone of therapy for AKI. This is based on the assumption that decrease in kidney blood flow is a major driver of AKI(29), However, three key observations challenge this hypothesis. First, even though fluid administration may increase cardiac output and renal blood flow, it does not lead to improvement in renal oxygen delivery or renal microvascular oxygenation(30, 31). Second, fluid bolus does not always increase urine output. In a prospective, multicenter, observational study including 5 ICUs where patients with oliguria received IV fluids, improvement in urine output was seen in only half of the patients(4). Third, kidney blood flow is not decreased in sepsis(32). Instead, the pathogenesis of AKI in critically ill patients includes a complex interplay of inflammatory mediators(33–35), microcirculatory dysfunction(36) and metabolic reprogramming(37, 38). These result in direct tubular injury, which is not reversible with administration of IV fluids.

IV fluid administration can lead to volume overload, often seen in AKI patients, and is associated with higher morbidity and mortality especially in AKI patients (39, 40). This excess fluid can cause interstitial edema(41, 42), impaired blood flow and organ dysfunctions. Fluid overload also damages the endothelial glycocalyx layer which is crucial for vascular homeostasis and permeability. Inflammatory mediators in sepsis themselves lead to glycocalyx degradation, a process which is exacerbated by IV fluid resuscitation(43). This damage increases local inflammation, tissue edema and further end organ injury including acute respiratory distress syndrome (ARDS) and AKI(44). Fluid overload is also a known risk-factor for development of intra-abdominal hypertension(45), which further contributes to worsening AKI (46). Thus, there is an increasing focus on minimizing the use of IV fluids in critically ill patients. A randomized study comparing conservative and usual fluid therapy in patients with ARDS found no difference in 60-day mortality(8). However, the conservative group had improved oxygenation index, lung injury score, lower plateau pressures, increased number of ventilator free days and ICU free days. Studies in patients with sepsis have also found a shorter duration of mechanical ventilation and a trend towards lower mortality for patients managed with conservative fluid strategy(47).

The REVERSE-AKI trial directly evaluated the role of restrictive fluid therapy for management of AKI(10). This multicenter, randomized, controlled study enrolled adult, critically ill patients with AKI not requiring dialysis. The trial aimed to attain a negative fluid balance in the intervention arm following randomization. By 24 hours, the restrictive fluid group had a cumulative fluid balance of -416mL vs 409mL in the usual care group (p<0.001). At 72 hours, this balance was -1080mL vs 61mL (p=0.03). The trial found that the restrictive fluid group had lower dialysis rates (13% vs 30%; p=0.04) and a trend towards lower duration of AKI (median 2 days vs 3 days; p=0.07). This trial demonstrated that restrictive fluid management in AKI is feasible and may improve outcomes. Building upon this, our study adds precision by identifying the subgroup of patients with AKI and sepsis most likely to benefit from fluid restriction.

Causal forests outperformed non-causal random forests in capturing HTEs, with higher AUTOC values in both cohorts. In the external validation cohort, an AUTOC of 0.15 indicates that patients prioritized by the model had, on average, a 15% greater probability of early AKI reversal with restrictive fluids compared with random treatment allocation. This clinically meaningful effect size showcases the potential of causal ML to uncover meaningful heterogeneity in treatment response that is typically obscured in average treatment effect analyses. Importantly, our policy tree stratified patients using routinely available clinical features such as respiratory rate, blood pressure, BUN and other bedside accessible measures, some of which may not traditionally guide fluid management decisions, making the proposed strategy both novel and practical for bedside application.. Our results also demonstrate the clinical value of adhering to model-guided recommendations. Among patients identified to benefit from restrictive fluids, those who received such care had significantly improved outcomes across all endpoints, whereas those not predicted to benefit showed no appreciable advantage from fluid restriction. This supports the central premise of treatment effect heterogeneity that not all patients benefit equally from one-size-fits-all protocols.

Despite these strengths, our study has limitations. First, although causal inference techniques aim to address confounding, residual bias from unmeasured variables cannot be entirely excluded. Second, we chose the cut-off of 500mL for restrictive fluids based prior literature suggesting 500mL is the typical amount of fluid given during a fluid challenge(3), and was also the amount given in restrictive fluid groups in prior studies(9, 48). This may, however, not capture all nuances of fluid management decisions and future work should focus on alternate fluid cutoffs. Third, we observed lower AUTOC values in the external validation cohort. This likely reflects expected challenges in transporting ITE models across international cohort with different case mixes, practice patterns, and underlying treatment heterogeneity. Nonetheless, the fact that causal forest continued to outperform non-causal model and yielded clinically consistent findings in the external cohort supports the robustness of this approach. Fourth, as our goal was to assess the impact of fluid volume rather than fluid type, we did not distinguish between different fluids administered. Future studies should evaluate the relative benefits of specific fluid types alongside restrictive strategies in this population. Finally, although our external validation supports generalizability, both datasets were from high-resource, academic settings. Future work should validate these findings in diverse clinical environments and prospective studies. Integration of causal ML into clinical workflows through decision support tools or trial enrichment strategies represents a promising direction for improving care in this high-risk population.

## CONCLUSION

In conclusion, we developed and validated a causal ML framework to identify critically ill patients with sepsis and AKI most likely to benefit from a restrictive fluid strategy. This personalized, data-driven approach holds promise for improving outcomes by incorporation into future studies and clinical workflows and should be evaluated in future prospective and pragmatic clinical trials.

## Data Availability

All data produced are available online at the Physionet website.

https://physionet.org/content/mimiciv/3.0/

https://physionet.org/content/sicdb/1.0.8/

## Funding

This work was supported by the National Institutes of Health (NIH) grants K08DK131286 awarded to A.S., R01DK108803, U01HG007278, U01HG009610, and U01DK116100 awarded to G.N.N. and T32DK007757 and TL1DK136048 awarded to W.O. The content is solely the responsibility of the authors and does not necessarily represent the views of the NIH.

## Competing interests

GNN is a founder of Renalytix, Pensieve, Verici and provides consultancy services to AstraZeneca, Reata, Renalytix, Siemens Healthineer and Variant Bio, serves a scientific advisory board member for Renalytix and Pensieve. He also has equity in Renalytix, Pensieve and Verici. LC is a consultant for Vifor Pharma INC and has received honorarium from Fresenius Medical Care. AS is a consultant for Roche Diagnostics Corporation. JAK reports receiving consulting fees from Astute Medical/bioMerieux, Astellas, Alexion, Chugai Pharma, Novartis, Mitsubishi Tenabe and GE Healthcare and is a Full-time employee of Spectral Medical. All remaining authors have declared no conflicts of interest.

## Data availability

Publicly available datasets were analyzed in this study. The dataset used in this study, MIMIC-IV, is available at https://physionet.org/content/mimiciv/, and the SICdb dataset is available at https://physionet.org/content/sicdb/.

## Code availability

Source code available upon reasonable request.

## Author contributions

WO and AS designed and conceptualized the study. WO, KT, ZA and HK performed data collection and analysis. WO and AS interpreted the data analyses. WO and AS wrote the first draft of the manuscript. All authors critically revised the manuscript for important intellectual content and read and approved the final version. All authors gave final approval for the version to be published. WO is first author, GNN and AS and co-senior authors.

## Notes

### Author Declarations

The study received approval from the Institutional Review Board at the Icahn School of Medicine at Mount Sinai (approval no. 19-00951).

### Summary of Updates

In this revised version, we emphasize the estimation of heterogeneous treatment effects using causal machine learning methods, evaluated through the Area Under the Targeting Operator Characteristic (AUTOC) metric.

