## Supplementary document for "Personalized Fluid Management in Patients with Sepsis and AKI: A Casual Machine Learning Approach"

**Supplementary Materials**

**Table of Contents**

| Contents | Page number |
| --- | --- |
| **Supplementary Methods** | 2 |
| **Supplementary Results** | 9 |
| **Supplementary Figure S1.** Cohort Inclusion and Exclusion Criteria. | 11 |
| **Supplementary Table S1.** TRIPOD+AI checklist | 12 |
| **Supplementary Table S2.** Comparison of additional baseline characteristics of patients identified by the policy tree as likely to benefit versus not benefit from restrictive fluids | 17 |
| **Supplementary Table S3.** Comparison of additional baseline characteristics of patients who actually received versus did not receive restrictive fluids within the subgroup predicted to benefit from restrictive fluids | 24 |
| **Supplementary Table S4.** Sensitivity analysis showing impact of restrictive fluid strategy among patients with congestive heart failure who were predicted to benefit from restrictive fluids | 32 |
| **Supplementary Table S5.** Sensitivity analysis showing impact of restrictive fluid strategy among patients with chronic kidney disease who were predicted to benefit from restrictive fluids | 33 |
| **Supplementary Table S6.** Sensitivity analysis showing impact of restrictive fluid strategy among patients who were predicted to benefit from restrictive fluids when admission creatinine was treated as baseline creatinine for all patients | 34 |

**SUPPLEMENTARY METHODS:**

**Determination of Baseline Creatinine for identification of AKI**

To determine baseline serum creatinine, we used a tiered approach(1-5). First, we identified the reference creatinine by calculating the median of all available serum creatinine measurements taken between 1 and 365 days prior to hospital admission. We then compared this median reference value to the first recorded creatinine from the current hospital encounter and selected the lower of the two as the baseline creatinine. If no prior creatinine values were available, we estimated the reference creatinine using the Modification of Diet in Renal Disease equation (6) by back-calculating serum creatinine assuming an estimated glomerular filtration rate (eGFR) of 75 ml/min/1.73 m². This estimated value was then compared to the first creatinine measured during the current admission, and the lower of the two was used. For patients with a known history of chronic kidney disease, we used the first admission creatinine instead of the MDRD-based estimate(4). Patients with a baseline creatinine ≥ 4.0 mg/dL were excluded from the analysis to avoid confounding due to advanced underlying renal disease(5, 7).

**Development of Causal ML Approach**

We used a novel, causal machine learning (ML) strategy to identify critically ill septic patients with AKI who would benefit from a restrictive fluid strategy. This casual ML approach employed a dual step methodology. In the first step, we used causal forest to estimate individual treatment effects (ITE) to ascertain the impact restrictive fluid strategy at the individual patient level(8). Subsequently, we applied the policy tree algorithm(9) to construct a decision tree(10) based on the ITEs estimated by the causal forest model.

**Step 1 - Causal Forest**: We utilized the causal forest(8) model to estimate ITEs within our development cohort. This method is a widely recognized quasi-experimental approach for estimating treatment effects using observational data. Unlike traditional predictive models that focus on minimizing prediction error, causal forests aim to estimate **conditional average treatment effects (CATEs)** at the individual level by constructing trees that **maximize variation in treatment effects** between subgroups. These CATEs serve as approximations of ITEs based on observed patient characteristics.

Mathematically, given a dataset $\left\{ X_{i},W_{i},Y_{i} \right\}_{i=1}^{n}$, where $X_{i}$ represents patient covariates, $W_{i}\in\left\{ 0,1 \right\}$ represents treatment indicator ($W_{i}=1$ if treated, $0$ otherwise), and $Y_{i}$ the observed outcomes, the causal forest estimates CATE ($\hat{\Gamma_{i}}$) (11, 12) as follows:

$\hat{\Gamma_{i}}=\hat{m}\left( X_{i},1 \right)-\hat{m}\left( X_{i},0 \right)+\frac{\left( W_{i}-\hat{e}\left( X_{i} \right) \right)}{\hat{e}\left( X_{i} \right)\left( 1-\hat{e}\left( X_{i} \right) \right)}\left( Y_{i}-\hat{m}\left( X_{i},W_{i} \right) \right)$	(1)

Here, $\hat{e}\left( x \right)=P\left( W_{i}=1 | X_{i}=x \right)$ represents the propensity score, indicating the probability of receiving restricted fluid intake therapy based on patient characteristics, and $\hat{m}\left( x,w \right)\mathbb{=E}\left[ Y_{i}(w)|X_{i}=x \right]$ represents outcome model, estimating the expected outcome under each treatment condition. This estimator combines the treatment effect predicted by the outcome model, $\hat{m}\left( X_{i},1 \right)-\hat{m}\left( X_{i},0 \right)$, with a bias correction term using inverse probability weighting $\frac{\left( W_{i}-\hat{e}\left( X_{i} \right) \right)}{\hat{e}\left( X_{i} \right)\left( 1-\hat{e}\left( X_{i} \right) \right)}$, which is derived from the propensity score to balance the treated and untreated groups by reweighting samples. The resulting CATE estimate is doubly robust as it remains consistent if either the propensity or the outcome model is mis-specified(13). Initially, the causal forest algorithm identifies optimal splits that maximize the expected difference in treatment effects $\hat{\Gamma_{i}}$ across subgroups. Following that, it estimates treatment effects by aggregating data from similar observations within the forest's leaves.

**Step 2 - Policy Tree Algorithm**: We then used the policy tree algorithm(9) to identify patients most likely to benefit, in terms of early AKI reversal, from a restrictive fluid strategy. This algorithm constructs a hierarchical model that categorizes patients based on similar ITE within the same treatment node and distinct ITE across different treatment nodes. The goal is to learn an optimal treatment policy $\pi\left( x \right)$ that assigns treatments based on patient covariates $x$, maximizing the expected outcome defined as:

$\pi^{*}\left( x \right)=\mathrm{argmax}_{\pi\in\Pi} \left[ \sum_{i=1}^{n} \Gamma_{i}\left( \pi\left( X_{i} \right) \right) \right]$	(2)

where $\Pi$ is the class of depth-$k$ decision trees, $\Gamma$ is a vector of unit-specific rewards for each action $1$ to $d$, and $\pi\left( X_{i} \right)$ maps from covariates $X_{i}$ to action.

The depth of the tree was determined through cross-validation within our development cohort. A depth of five was chosen as it provided sufficient stratification of treatment effects without overfitting or compromising usability. Overfitting was assessed based on differences in the proportion of primary outcomes. In this way, the policy tree translates ITEs into an interpretable decision framework to guide treatment selection.

The resulting policy tree can be interpreted by following a top-down sequence of binary decision rules based on patient covariates. At each internal node, the algorithm applies a threshold-based split, directing patients down different branches based on whether the condition is met. This recursive partitioning continues until a terminal node (leaf) is reached, which assigns a treatment recommendation based on the subgroup’s estimated treatment benefit. In this way, the policy tree translates ITE estimates into an interpretable sequence of clinical decision rules that guide treatment selection.

**Statistical Analysis**

We expressed continuous features as mean and standard deviation, while categorical features as proportions. We examined the differences across groups using bivariate analyses, including analysis of variance (ANOVA) for continuous variables and chi-square tests for categorical variables, with a P value of < .05 considered statistically significant across all statistical tests.

To evaluate how effectively our causal machine learning approach captured heterogeneous treatment effects (HTEs), we computed the **area under the targeting operator characteristic curve (AUTOC)** (12). AUTOC quantifies how effectively a model ranks patients by their expected benefit from treatment (ITE). A higher AUTOC value indicates that the model more accurately prioritized patients who truly benefited from the intervention.

Following established methodology, we estimated the **average treatment effect (ATE)** using **augmented inverse probability weighting (AIPW),** a doubly robust method that combines inverse probability weighting and outcome regression to provide unbiased population-level treatment effect estimates(14, 15).

The AUTOC is derived from the targeting operating characteristic (TOC) curve. This curve is constructed by ranking patients by their estimated conditional average treatment effects (CATEs), which serve as proxies for ITEs(16). For a given proportion of patients ($\alpha$, ranging from 0 to 1), the TOC measures the additional treatment effect gained by treating only the top $\alpha$-fraction of patients compared to treating all patients. Mathematically$:$

$\text{TOC}\left( \alpha\right)=\frac{1}{\left| S_{\alpha} \right|}\sum_{i\in S_{\alpha}} \tau\left( X_{i} \right)\mathbb{-E}\left( \tau\left( X \right) \right)$	(3)

where, $X_{i}$ represents patient characteristics, $\hat{\tau}\left( X \right)$the estimated conditional average treatment effect (CATE), $\tau\left( X \right)$ the average treatment effect for the entire population (ATE), and $S_{\alpha}$ is the group consisting of the top fraction $\alpha$ of patients with the highest estimated CATEs. The AUTOC is then computed as the area under this curve across all values of α from 0 to 1:

$\text{AUTOC}=\int_{0}^{1} \text{TOC}\left( \alpha\right)d\alpha$.	(4)

We trained both **causal forest** and **random forest** models on the development cohort (MIMIC-IV) to estimate ITEs. The random forest served as the non-causal ML comparator model to evaluate if causal ML model does indeed better capture HTEs. The causal forest and random forest models trained on MIMIC-IV were then validated in SiCdb to generate patient-level treatment effect estimates. We then computed AUTOC for each model in each dataset, using the dataset-specific ATE from AIPW as the baseline. Finally, we compared the AUTOC values between causal forest and random forest within each dataset using a two-sample t-test to assess whether the causal model more effectively captured meaningful treatment effect heterogeneity

To examine the clinical impact of adhering to personalized restrictive fluid recommendations derived from the policy tree we compared unadjusted event rates for early AKI reversal, sustained AKI reversal, and MAKE30 between patients who adhered to restrictive fluid recommendations and those who did not, using proportions and chi-square tests. Subsequently, we quantified these associations using logistic regression models to estimate odds ratios (OR) and 95% confidence intervals (CI), adjusting for age, sex, race, reference serum creatinine, SOFA score at the time of sepsis onset, AKI stage at the time of AKI onset, and net fluid balance between ICU admission and AKI onset for each outcome.

**Sensitivity Analyses**

To evaluate the robustness of our findings, we performed two sensitivity analyses. First we evaluated the performance of policy tree in patients with congestive heart failure (CHF) and chronic kidney disease (CKD). While CKD was available in both datasets, CHF status was only available in the development cohort. Second, we evaluated the performance of policy tree by redefining the reference creatinine as the first admission creatinine among those without a known pre-admission value, while retaining the observed pre-admission baseline when available.

**SUPPLEMENTARY RESULTS:**

**Patients with CHF:** Information about CHF diagnosis was not available for patients in external validation cohort. In the development cohort, 7,477 (64.2%) patients had CHF. Among those with CHF who were identified by policy tree as likely to benefit from a restrictive fluid strategy, patients who actually received restrictive fluids had substantially higher rates of early AKI reversal (57.8% vs 32.7%; p<0.001), sustained AKI reversal (23.5% vs 13.5%; p<0.001), and lower rates of MAKE30 (26.3% vs 32.4%; p=0.003) compared with those who did not receive restrictive fluids (Supplementary Table S4).

**Patients with CKD:** 2,364 patients (20.3 %) in development cohort and 303 (15.7%) patients in external validation cohort had CKD. In the development cohort, among those predicted to benefit from restrictive fluids, those who received restrictive fluids had higher rates of early AKI reversal (50.0% vs 27.5%; p<0.001), sustained AKI reversal (21.8% vs 12.1%; p<0.001) and lower rates of MAKE30 (21.3% vs 32.5%; p=0.003).

In the external validation cohort, patients predicted to benefit from restrictive fluids who received restrictive fluids similarly demonstrated higher rates of early AKI reversal (33.3% vs 19.0%, p=0.4), sustained AKI reversal (25.0% vs 10.3%, p=0.3) and lower rates of MAKE30 (25.0% vs 48.3%, p=0.2), though they did not reach statistical significant likely due to small sample size (Supplementary Table S5).

Thus, even though the model input did not include comorbidities, our model generalizes well to patients with CHF and CKD.

**Baseline Creatinine Sensitivity Analysis**

The MDRD equation was used to back-calculate the baseline creatinine for 1,687 (14.5%) patients in the development cohort (MIMIC-IV) and 902 (46.7%) patients in the external validation cohort (SICdb). To ensure that our results were not driven by assumptions around baseline creatinine, we performed a sensitivity analysis redefining baseline creatinine as the first admission creatinine in patients without a known pre-admission value, while retaining the observed pre-admission baseline value when available. Using this approach in MIMIC-IV, we identified 11,541 patients with sepsis and AKI, of which 5,075 (44.0%) were identified to benefit from restrictive fluids but only 807 (15.9%) received them. In SICdb, 1,841 patients met inclusion criteria with 1,271 (69.0%) predicted to benefit from restrictive fluid strategy. Of them, only 71 (5.6%) received restrictive fluids.

In this sensitivity analysis, the causal forest continued to outperform the random forest in identifying HTE of restrictive fluids for early AKI reversal in both development cohort (AUTOC for causal forest was 0.46; 95% CI: 0.43-0.49 vs 0.34; 95% CI 0.31-0.37 for random forest) and external validation cohort (AUTOC for causal forest was 0.08; 95% CI: 0.01-0.16 vs -0.05; 95% CI -0.15-0.06 for random forest). In the development cohort, among patients predicted to benefit from restrictive fluids, those who received restrictive fluids had higher rates of early AKI reversal (53.4% vs. 37.2%; p<0.001), sustained AKI reversal (23.4% vs.17.6 %; p<0.001), and lower rates of MAKE30 (21.3% vs. 26.4%; p=0.003). Similarly in external validation cohort, among patients recommended to benefit from restrictive fluids, those who received them had significantly higher rates of early AKI reversal (56.3% vs. 36.0%; p<0.001), sustained AKI reversal (33.8% vs. 19.8%; p=0.007), and lower rates of MAKE30 (16.9% vs. 30.4%; p=0.02). (Supplementary Table S6).”

**Supplementary Figure**

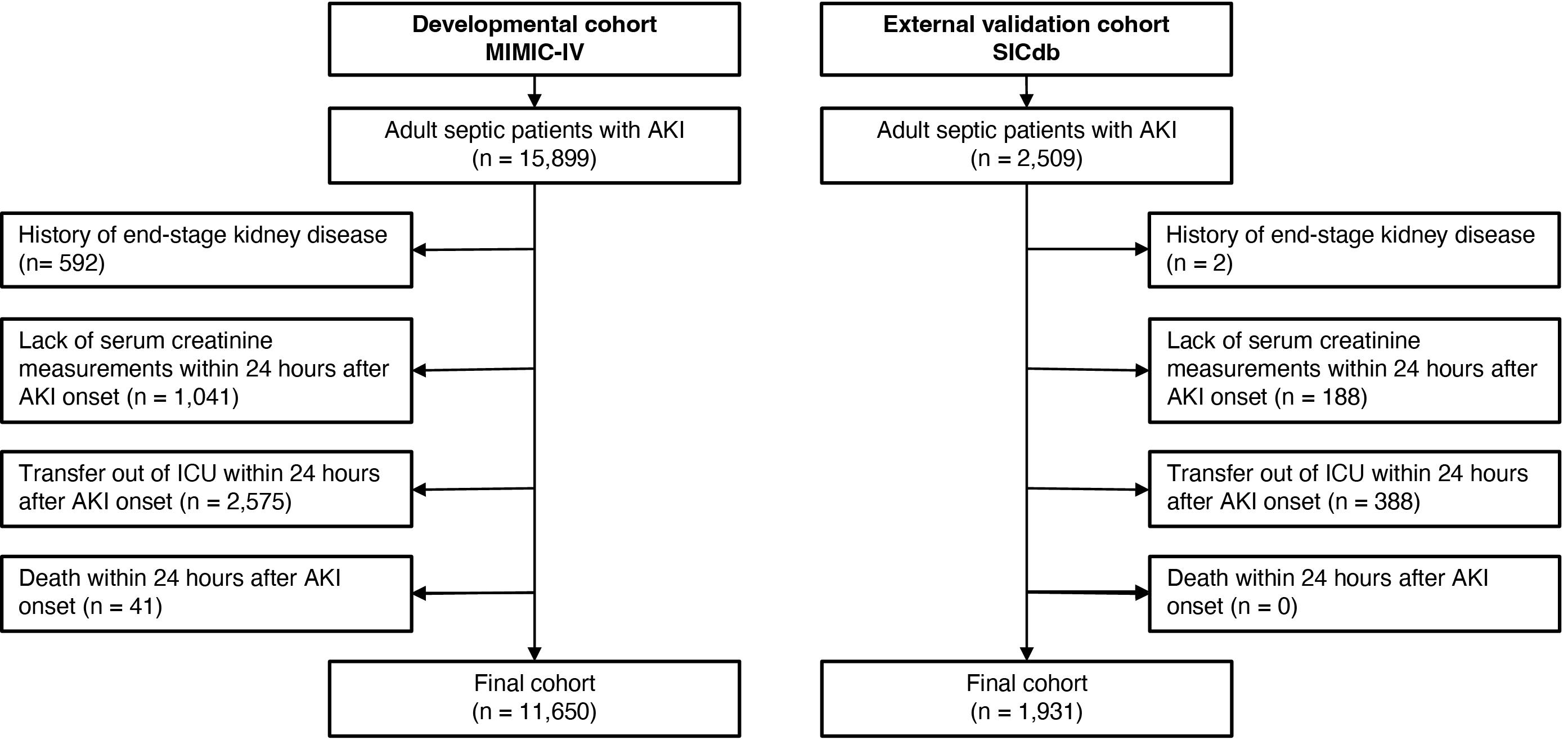

**Supplementary Figure S1.** Cohort Inclusion and Exclusion Criteria.

**Supplementary Tables**

**Supplementary Table S1.** TRIPOD+AI checklist

| **Section/Topic Item ^Development^ Checklist item / evaluation**^1^ | | | | **Reported on page** |
| --- | --- | --- | --- | --- |
| **TITLE** | | | |  |
| *Title* | 1 | D;E | Identify the study as developing or evaluating the performance of a multivariable prediction model, the target population, and the outcome to be predicted | 1 |
| **ABSTRACT** | | | | |
| *Abstract* | 2 | D;E | See TRIPOD+AI for Abstracts checklist | 4 |
| **INTRODUCTION** | | | | |
| *Background* | 3a | D;E | Explain the healthcare context (including whether diagnostic or prognostic) and rationale for developing or evaluating the prediction model, including references to existing models | 7 |
|  | 3b | D;E | Describe the target population and the intended purpose of the prediction model in the context of the care pathway, including its intended users (e.g., healthcare professionals, patients, public) | 7 |
|  | 3c | D;E | Describe any known health inequalities between sociodemographic groups | NA |
| *Objectives* | 4 | D;E | Specify the study objectives, including whether the study describes the development or validation of a prediction model (or both) | 8 |
| **METHODS** | | | | |
| *Data* | 5a | D;E | Describe the sources of data separately for the development and evaluation datasets (e.g., randomised trial, cohort, routine care or registry data), the rationale for using these data, and representativeness of the data | 8 |
|  | 5b | D;E | Specify the dates of the collected participant data, including start and end of participant accrual; and, if applicable, end of follow-up | 8 |
| *Participants* | 6a | D;E | Specify key elements of the study setting (e.g., primary care, secondary care, general population) including the number and location of centres | 8 |
|  | 6b | D;E | Describe the eligibility criteria for study participants | 8 |
|  | 6c | D;E | Give details of any treatments received, and how they were handled during model development or evaluation, if relevant | 9 |
| *Data preparation* | 7 | D;E | Describe any data pre-processing and quality checking, including whether this was similar across relevant sociodemographic groups | 10 |
| *Outcome* | 8a | D;E | Clearly define the outcome that is being predicted and the time horizon, including how and when assessed, the rationale for choosing this outcome, and whether the method of outcome assessment is consistent across sociodemographic groups | 9 |
|  | 8b | D;E | If outcome assessment requires subjective interpretation, describe the qualifications and demographic characteristics of the outcome assessors | NA |
|  | 8c | D;E | Report any actions to blind assessment of the outcome to be predicted | NA |
| *Predictors* | 9a | D | Describe the choice of initial predictors (e.g., literature, previous models, all available predictors) and any pre-selection of predictors before model building | 10 |
|  | 9b | D;E | Clearly define all predictors, including how and when they were measured (and any actions to blind assessment of predictors for the outcome and other predictors) | 10 |
|  | 9c | D;E | If predictor measurement requires subjective interpretation, describe the qualifications and demographic characteristics of the predictor assessors | NA |
| *Sample size* | 10 | D;E | Explain how the study size was arrived at (separately for development and evaluation), and justify that the study size was sufficient to answer the research question. Include details of any sample size calculation | 9 |
| *Missing data* | 11 | D;E | Describe how missing data were handled. Provide reasons for omitting any data | 10 |
| *Analytical methods* | 12a | D | Describe how the data were used (e.g., for development and evaluation of model performance) in the analysis, including whether the data were partitioned, considering any sample size requirements | 10 |
|  | 12b | D | Depending on the type of model, describe how predictors were handled in the analyses (functional form, rescaling, transformation, or any standardisation). | 10 |
|  | 12c | D | Specify the type of model, rationale^2^, all model-building steps, including any hyperparameter tuning, and method for internal validation | 10, Supp. 2 |
|  | 12d | D;E | Describe if and how any heterogeneity in estimates of model parameter values and model performance was handled and quantified across clusters (e.g., hospitals, countries). See TRIPOD-Cluster for additional considerations^3^ | NA |
|  | 12e | D;E | Specify all measures and plots used (and their rationale) to evaluate model performance (e.g., discrimination, calibration, clinical utility) and, if relevant, to compare multiple models | 11 |
|  | 12f | E | Describe any model updating (e.g., recalibration) arising from the model evaluation, either overall or for particular sociodemographic groups or settings | NA |
|  | 12g | E | For model evaluation, describe how the model predictions were calculated (e.g., formula, code, object, application programming interface) | 11 |
| *Class imbalance* | 13 | D;E | If class imbalance methods were used, state why and how this was done, and any subsequent methods to recalibrate the model or the model predictions | NA |
| *Fairness* | 14 | D;E | Describe any approaches that were used to address model fairness and their rationale | NA |
| *Model output* | 15 | D | Specify the output of the prediction model (e.g., probabilities, classification). Provide details and rationale for any classification and how the thresholds were identified | 11, Supp. 4 |
| *Training versus evaluation* | 16 | D;E | Identify any differences between the development and evaluation data in healthcare setting, eligibility criteria, outcome, and predictors | 12 |
| *Ethical approval* | 17 | D;E | Name the institutional research board or ethics committee that approved the study and describe the participant-informed consent or the ethics committee waiver of informed consent | 12 |
| *Funding* | 18a | D;E | Give the source of funding and the role of the funders for the present study | 3 |
| *Conflicts of interest* | 18b | D;E | Declare any conflicts of interest and financial disclosures for all authors | 3 |
| *Protocol* | 18c | D;E | Indicate where the study protocol can be accessed or state that a protocol was not prepared | NA |
| *Registration* | 18d | D;E | Provide registration information for the study, including register name and registration number, or state that the study was not registered | NA |
| *Data sharing* | 18e | D;E | Provide details of the availability of the study data | 19 |
| *Code sharing* | 18f | D;E | Provide details of the availability of the analytical code^4^ | 19 |
| **PATIENT & PUBLIC INVOLVEMENT** | | | | |
| *Patient & Public Involvement* | 19 | D;E | Provide details of any patient and public involvement during the design, conduct, reporting, interpretation, or dissemination of the study or state no involvement. | 9 |
| **RESULTS** | | | | |
| *Participants* | 20a | D;E | Describe the flow of participants through the study, including the number of participants with and without the outcome and, if applicable, a summary of the follow-up time. A diagram may be helpful. | 12 |
|  | 20b | D;E | Report the characteristics overall and, where applicable, for each data source or setting, including the key dates, key predictors (including demographics), treatments received, sample size, number of outcome events, follow-up time, and amount of missing data. A table may be helpful. Report any differences across key demographic groups. | 12 |
|  | 20c | E | For model evaluation, show a comparison with the development data of the distribution of important predictors (demographics, predictors, and outcome). | 13 |
| *Model development* | 21 | D;E | Specify the number of participants and outcome events in each analysis (e.g., for model development, hyperparameter tuning, model evaluation) | 12 |
| *Model specification* | 22 | D | Provide details of the full prediction model (e.g., formula, code, object, application programming interface) to allow predictions in new individuals and to enable third-party evaluation and implementation, including any restrictions to access or re-use (e.g., freely available, proprietary)^5^ | 10, Supp. 2 |
| *Model performance* | 23a | D;E | Report model performance estimates with confidence intervals, including for any key subgroups (e.g., sociodemographic). Consider plots to aid presentation. | 12 |
|  | 23b | D;E | If examined, report results of any heterogeneity in model performance across clusters. See TRIPOD Cluster for additional details^3^. | NA |
| *Model updating* | 24 | E | Report the results from any model updating, including the updated model and subsequent performance | NA |
| **DISCUSSION** | | | | |
| *Interpretation* | 25 | D;E | Give an overall interpretation of the main results, including issues of fairness in the context of the objectives and previous studies | 15 |
| *Limitations* | 26 | D;E | Discuss any limitations of the study (such as a non-representative sample, sample size, overfitting, missing data) and their effects on any biases, statistical uncertainty, and generalizability | 18 |
| *Usability of the model in the context of current care* | 27a | D | Describe how poor quality or unavailable input data (e.g., predictor values) should be assessed and handled when implementing the prediction model | 18 |
|  | 27b | D | Specify whether users will be required to interact in the handling of the input data or use of the model, and what level of expertise is required of users | NA |
|  | 27c | D;E | Discuss any next steps for future research, with a specific view to applicability and generalizability of the model | 18 |

**Supplementary Table S2.** Comparison of additional baseline characteristics of patients identified by the policy tree as likely to benefit versus not benefit from restrictive fluids

|  | **Development cohort (MIMIC-IV)** | | | | **External validation cohort (SICdb)** | | | |
| --- | --- | --- | --- | --- | --- | --- | --- | --- |
|  | **Identified to have no benefit from restrictive fluid strategy**  **(N= 6,526)** | **Identified to benefit from restrictive fluid strategy**  **(N= 5,124)** | **Total**  **(N= 11,650)** | **p value** | **Identified to have no benefit from restrictive fluid strategy**  **(N= 600)** | **Identified to benefit from restrictive fluid strategy**  **(N= 1,331)** | **Total**  **(N= 1,931)** | p value |
| **Following restrictive fluid strategy, n (%)** | 1094 (16.8%) | 798 (15.6%) | 1892 (16.2%) | 0.08 | 53 (8.8%) | 76 (5.7%) | 129 (6.7%) | 0.01 |
| **The time (hour) between ICU admission and AKI onset, (mean±SD)** | 14.9 ± 10.3 | 14.5 ± 10.2 | 14.7 ± 10.2 | 0.02 | 11.0 ± 7.9 | 13.3 ± 9.9 | 12.6 ± 9.4 | < 0.001 |
| **Latest heart rate^A^, (mean±SD)** | 86.5 ± 19.1 | 88.0 ± 21.0 | 87.2 ± 20.0 | < 0.001 | 80.0 ± 18.3 | 86.4 ± 19.4 | 84.4 ± 19.3 | < 0.001 |
| **Maximum heart rate^A^, (mean±SD)** | 103.4 ± 21.6 | 104.2 ± 23.5 | 103.8 ± 22.5 | 0.08 | 114.8 ± 23.4 | 136.7 ± 42.0 | 129.9 ± 38.6 | < 0.001 |
| **Minimum heart rate^A^, (mean±SD)** | 76.1 ± 17.7 | 76.2 ± 19.2 | 76.2 ± 18.4 | 0.6 | 61.6 ± 16.8 | 61.7 ± 17.5 | 61.7 ± 17.3 | 0.9 |
| **Latest SBP^A^, mmHg, (mean±SD)** | 113.5 ± 20.2 | 112.2 ± 21.0 | 112.9 ± 20.6 | < 0.001 | 108.6 ± 20.1 | 104.7 ± 20.6 | 105.9 ± 20.5 | < 0.001 |
| **Maximum SBP^A^, mmHg, (mean±SD)** | 142.8 ± 24.9 | 142.1 ± 25.0 | 142.5 ± 24.9 | 0.1 | 205.8 ± 61.2 | 217.7 ± 65.9 | 214.0 ± 64.7 | < 0.001 |
| **Minimum SBP^A^, mmHg, (mean±SD)** | 92.5 ± 17.0 | 88.4 ± 18.8 | 90.7 ± 18.0 | < 0.001 | 65.2 ± 26.8 | 54.8 ± 22.1 | 58.0 ± 24.1 | < 0.001 |
| **Latest DBP^A^, mmHg, (mean±SD)** | 60.5 ± 14.2 | 60.1 ± 14.4 | 60.3 ± 14.3 | 0.2 | 54.7 ± 11.2 | 53.7 ± 10.3 | 54.0 ± 10.6 | 0.06 |
| **Maximum DBP^A^, mmHg, (mean±SD)** | 83.7 ± 20.4 | 83.4 ± 21.2 | 83.6 ± 20.7 | 0.5 | 119.3 ± 56.8 | 130.5 ± 65.4 | 127.0 ± 63.1 | < 0.001 |
| **Minimum DBP^A^, mmHg, (mean±SD)** | 47.9 ± 11.9 | 46.3 ± 12.3 | 47.2 ± 12.1 | < 0.001 | 29.0 ± 14.6 | 24.7 ± 13.2 | 26.0 ± 13.8 | < 0.001 |
| **Latest MBP^A^, mmHg, (mean±SD)** | 75.6 ± 15.0 | 75.2 ± 15.1 | 75.4 ± 15.0 | 0.1 | 73.1 ± 15.2 | 70.9 ± 12.6 | 71.6 ± 13.5 | < 0.001 |
| **Maximum MBP^A^, mmHg, (mean±SD)** | 100.8 ± 25.1 | 101.0 ± 26.7 | 100.9 ± 25.8 | 0.6 | 163.0 ± 58.2 | 176.0 ± 60.8 | 172.0 ± 60.3 | < 0.001 |
| **Minimum MBP^A^, mmHg, (mean±SD)** | 60.4 ± 14.0 | 58.0 ± 14.8 | 59.3 ± 14.4 | < 0.001 | 36.3 ± 20.3 | 28.9 ± 18.2 | 31.2 ± 19.2 | < 0.001 |
| **Latest respiratory rate^A^, (mean±SD)** | 19.8 ± 5.8 | 20.7 ± 5.8 | 20.2 ± 5.8 | < 0.001 | 16.7 ± 5.7 | 15.9 ± 5.4 | 16.1 ± 5.5 | 0.005 |
| **Maximum respiratory rate^A^, (mean±SD)** | 27.2 ± 7.1 | 27.5 ± 6.9 | 27.3 ± 7.0 | 0.1 | 28.8 ± 12.5 | 30.4 ± 13.5 | 29.9 ± 13.2 | 0.03 |
| **Minimum respiratory rate^A^, (mean±SD)** | 14.1 ± 4.6 | 14.4 ± 4.7 | 14.2 ± 4.6 | 0.003 | 10.2 ± 4.4 | 8.3 ± 4.7 | 8.9 ± 4.7 | < 0.001 |
| **Latest temperature^A^, C, (mean±SD)** | 36.9 ± 0.8 | 36.9 ± 0.8 | 36.9 ± 0.8 | 0.7 | 36.4 ± 2.5 | 36.7 ± 2.1 | 36.6 ± 2.2 | 0.008 |
| **Maximum temperature^A^, C, (mean±SD)** | 37.3 ± 0.9 | 37.3 ± 0.9 | 37.3 ± 0.9 | 0.1 | 37.4 ± 1.3 | 37.5 ± 1.1 | 37.5 ± 1.2 | 0.03 |
| **Minimum temperature^A^, C, (mean±SD)** | 36.4 ± 0.8 | 36.3 ± 0.9 | 36.4 ± 0.9 | < 0.001 | 31.6 ± 5.3 | 31.7 ± 5.1 | 31.7 ± 5.2 | 0.6 |
| **Latest SpO2^A^, %, (mean±SD)** | 97.0 ± 3.1 | 96.7 ± 3.6 | 96.9 ± 3.3 | < 0.001 | 96.0 ± 3.3 | 96.2 ± 4.0 | 96.1 ± 3.8 | 0.4 |
| **Maximum SpO2^A^, %, (mean±SD)** | 99.3 ± 1.5 | 99.2 ± 1.9 | 99.3 ± 1.7 | 0.04 | 99.5 ± 1.1 | 99.7 ± 1.2 | 99.7 ± 1.2 | < 0.001 |
| **Minimum SpO2^A^, %, (mean±SD)** | 92.6 ± 6.3 | 92.2 ± 6.7 | 92.4 ± 6.5 | < 0.001 | 82.7 ± 10.7 | 80.6 ± 11.7 | 81.3 ± 11.5 | < 0.001 |
| **Weight^A^, Kg, (mean±SD)** | 84.7 ± 23.2 | 85.2 ± 25.8 | 84.9 ± 24.3 | 0.3 | 82.3 ± 22.7 | 79.9 ± 22.6 | 80.7 ± 22.7 | 0.04 |
| **Latest hematocrit^A^, %, (mean±SD)** | 32.3 ± 6.6 | 32.0 ± 6.3 | 32.2 ± 6.4 | 0.01 | 34.5 ± 7.1 | 30.4 ± 5.8 | 31.6 ± 6.5 | < 0.001 |
| **Maximum hematocrit^A^, %, (mean±SD)** | 34.2 ± 6.7 | 34.1 ± 6.3 | 34.1 ± 6.6 | 0.2 | 35.9 ± 7.2 | 32.5 ± 6.2 | 33.5 ± 6.7 | < 0.001 |
| **Minimum hematocrit^A^, %, (mean±SD)** | 31.2 ± 7.1 | 30.7 ± 6.8 | 31.0 ± 7.0 | 0.001 | 34.1 ± 7.2 | 29.7 ± 6.1 | 31.0 ± 6.8 | < 0.001 |
| **Latest hemoglobin^A^, g/dL, (mean±SD)** | 10.6 ± 2.2 | 10.6 ± 2.1 | 10.6 ± 2.2 | 0.6 | 11.6 ± 2.3 | 10.3 ± 2.0 | 10.7 ± 2.2 | < 0.001 |
| **Maximum hemoglobin^A^, g/dL, (mean±SD)** | 11.1 ± 2.3 | 11.2 ± 2.1 | 11.2 ± 2.2 | 0.6 | 12.1 ± 2.4 | 11.1 ± 2.2 | 11.4 ± 2.3 | < 0.001 |
| **Minimum hemoglobin^A^, g/dL, (mean±SD)** | 10.2 ± 2.4 | 10.1 ± 2.3 | 10.2 ± 2.3 | 0.1 | 11.5 ± 2.4 | 10.1 ± 2.1 | 10.5 ± 2.3 | < 0.001 |
| **Latest MCH^A^, pg, (mean±SD)** | 29.9 ± 2.8 | 30.1 ± 2.7 | 30.0 ± 2.8 | < 0.001 | 29.8 ± 2.6 | 30.2 ± 2.4 | 30.1 ± 2.5 | < 0.001 |
| **Maximum MCH^A^, pg, (mean±SD)** | 30.1 ± 2.9 | 30.4 ± 2.8 | 30.2 ± 2.9 | < 0.001 | 29.9 ± 2.7 | 30.4 ± 2.5 | 30.3 ± 2.5 | < 0.001 |
| **Minimum MCH^A^, pg, (mean±SD)** | 29.7 ± 2.8 | 29.9 ± 2.8 | 29.8 ± 2.8 | < 0.001 | 29.7 ± 2.7 | 30.1 ± 2.5 | 30.0 ± 2.5 | 0.003 |
| **Latest MCHC^A^, g/L, (mean±SD)** | 32.7 ± 1.7 | 32.9 ± 1.8 | 32.8 ± 1.8 | < 0.001 | 33.7 ± 1.5 | 34.0 ± 1.5 | 33.9 ± 1.5 | < 0.001 |
| **Maximum MCHC^A^, g/L, (mean±SD)** | 32.9 ± 1.7 | 33.2 ± 1.9 | 33.0 ± 1.8 | < 0.001 | 34.0 ± 1.6 | 34.3 ± 1.5 | 34.2 ± 1.5 | < 0.001 |
| **Minimum MCHC^A^, g/L, (mean±SD)** | 32.3 ± 1.7 | 32.5 ± 1.7 | 32.4 ± 1.7 | < 0.001 | 33.6 ± 1.5 | 33.8 ± 1.5 | 33.7 ± 1.5 | 0.03 |
| **Latest MCV^A^, fL, (mean±SD)** | 91.6 ± 7.6 | 91.6 ± 7.3 | 91.6 ± 7.5 | 0.9 | 88.3 ± 6.6 | 88.9 ± 6.3 | 88.8 ± 6.4 | 0.06 |
| **Maximum MCV^A^, fL, (mean±SD)** | 92.5 ± 7.8 | 92.6 ± 7.4 | 92.5 ± 7.6 | 0.4 | 88.6 ± 6.6 | 89.5 ± 6.4 | 89.2 ± 6.5 | 0.007 |
| **Minimum MCV^A^, fL, (mean±SD)** | 91.1 ± 7.6 | 91.0 ± 7.3 | 91.1 ± 7.5 | 0.8 | 87.9 ± 6.6 | 88.4 ± 6.3 | 88.2 ± 6.4 | 0.1 |
| **Latest platelet^A^, K/uL, (mean±SD)** | 195.8 ± 109.7 | 200.1 ± 114.0 | 197.7 ± 111.7 | 0.05 | 239.9 ± 106.0 | 199.7 ± 101.7 | 212.0 ± 104.7 | < 0.001 |
| **Maximum platelet^A^, K/uL, (mean±SD)** | 214.3 ± 116.5 | 219.7 ± 119.8 | 216.7 ± 117.9 | 0.02 | 252.2 ± 106.6 | 221.1 ± 108.5 | 230.6 ± 108.8 | < 0.001 |
| **Minimum platelet^A^, K/uL, (mean±SD)** | 186.6 ± 106.5 | 190.1 ± 112.9 | 188.1 ± 109.4 | 0.1 | 233.8 ± 102.0 | 193.5 ± 101.1 | 205.7 ± 103.0 | < 0.001 |
| **Latest RBC^A^, Count m/uL, (mean±SD)** | 3.6 ± 0.8 | 3.5 ± 0.7 | 3.5 ± 0.8 | 0.004 | 3.9 ± 0.9 | 3.4 ± 0.6 | 3.6 ± 0.8 | < 0.001 |
| **Maximum RBC^A^ Count, m/uL, (mean±SD)** | 3.8 ± 0.8 | 3.7 ± 0.7 | 3.7 ± 0.8 | 0.07 | 4.1 ± 0.9 | 3.7 ± 0.7 | 3.8 ± 0.8 | < 0.001 |
| **Minimum RBC^A^ Count, m/uL, (mean±SD)** | 3.4 ± 0.8 | 3.4 ± 0.8 | 3.4 ± 0.8 | < 0.001 | 3.9 ± 0.9 | 3.3 ± 0.7 | 3.5 ± 0.8 | < 0.001 |
| **Latest WBC^A^, K/uL, (mean±SD)** | 13.8 ± 11.0 | 13.7 ± 9.7 | 13.8 ± 10.4 | 0.7 | 13.4 ± 7.4 | 12.5 ± 7.2 | 12.8 ± 7.3 | 0.02 |
| **Maximum WBC^A^, K/uL, (mean±SD)** | 15.2 ± 11.8 | 15.2 ± 10.2 | 15.2 ± 11.1 | 0.8 | 14.4 ± 7.5 | 13.9 ± 7.5 | 14.0 ± 7.5 | 0.2 |
| **Minimum WBC^A^, K/uL, (mean±SD)** | 12.2 ± 10.0 | 12.1 ± 9.2 | 12.2 ± 9.7 | 0.4 | 12.5 ± 7.3 | 11.1 ± 6.6 | 11.5 ± 6.8 | < 0.001 |
| **Latest anion gap^A^, mEq/L, (mean±SD)** | 14.8 ± 4.6 | 15.0 ± 4.6 | 14.9 ± 4.6 | 0.06 | 15.0 ± 3.9 | 13.9 ± 4.3 | 14.2 ± 4.2 | < 0.001 |
| **Maximum anion gap^A^, mEq/L, (mean±SD)** | 16.3 ± 4.9 | 16.4 ± 4.8 | 16.3 ± 4.9 | 0.1 | 17.4 ± 4.2 | 16.9 ± 4.4 | 17.1 ± 4.3 | 0.09 |
| **Minimum anion gap^A^, mEq/L, (mean±SD)** | 13.9 ± 4.4 | 14.0 ± 4.5 | 14.0 ± 4.5 | 0.4 | 13.8 ± 4.0 | 12.1 ± 4.9 | 12.6 ± 4.8 | < 0.001 |
| **Latest bicarbonate^A^, mEq/L, (mean±SD)** | 22.4 ± 5.2 | 22.1 ± 5.1 | 22.3 ± 5.1 | 0.02 | 24.5 ± 4.4 | 24.0 ± 4.4 | 24.1 ± 4.4 | 0.02 |
| **Maximum bicarbonate^A^, mEq/L, (mean±SD)** | 23.4 ± 5.3 | 23.2 ± 5.1 | 23.3 ± 5.2 | 0.2 | 25.8 ± 4.6 | 25.8 ± 4.7 | 25.8 ± 4.6 | 0.8 |
| **Minimum bicarbonate^A^, mEq/L, (mean±SD)** | 21.3 ± 5.3 | 21.1 ± 5.2 | 21.2 ± 5.2 | 0.04 | 21.9 ± 4.5 | 20.9 ± 4.4 | 21.2 ± 4.5 | < 0.001 |
| **Latest BUN^A^, mg/dL, (mean±SD)** | 31.5 ± 24.9 | 31.3 ± 24.1 | 31.4 ± 24.5 | 0.6 | 27.9 ± 21.1 | 25.9 ± 20.8 | 26.5 ± 20.9 | 0.06 |
| **Maximum BUN^A^, mg/dL, (mean±SD)** | 32.9 ± 25.3 | 32.6 ± 24.5 | 32.8 ± 24.9 | 0.6 | 28.7 ± 21.3 | 26.9 ± 21.3 | 27.4 ± 21.3 | 0.1 |
| **Minimum BUN^A^, mg/dL, (mean±SD)** | 29.9 ± 24.6 | 29.8 ± 23.9 | 29.9 ± 24.3 | 0.7 | 26.9 ± 20.9 | 24.9 ± 20.9 | 25.5 ± 20.9 | 0.07 |
| **Latest calcium^A^, mg/dL, (mean±SD)** | 8.2 ± 0.8 | 8.2 ± 0.9 | 8.2 ± 0.9 | 0.03 | 8.6 ± 0.7 | 8.4 ± 0.6 | 8.5 ± 0.7 | < 0.001 |
| **Maximum calcium^A^, mg/dL, (mean±SD)** | 8.4 ± 0.9 | 8.4 ± 0.9 | 8.4 ± 0.9 | 0.1 | 8.7 ± 0.7 | 8.6 ± 0.7 | 8.7 ± 0.7 | < 0.001 |
| **Minimum calcium^A^, mg/dL, (mean±SD)** | 8.1 ± 0.9 | 8.0 ± 0.9 | 8.0 ± 0.9 | 0.002 | 8.5 ± 0.7 | 8.4 ± 0.6 | 8.4 ± 0.7 | < 0.001 |
| **Latest chloride^A^, mEq/L, (mean±SD)** | 104.4 ± 6.8 | 104.1 ± 7.0 | 104.3 ± 6.9 | 0.05 | 99.7 ± 8.9 | 101.5 ± 6.2 | 100.9 ± 7.2 | 0.001 |
| **Maximum chloride^A^, mEq/L, (mean±SD)** | 105.4 ± 7.1 | 105.2 ± 7.3 | 105.3 ± 7.2 | 0.1 | 100.2 ± 6.4 | 101.6 ± 6.2 | 101.1 ± 6.3 | 0.006 |
| **Minimum chloride^A^, mEq/L, (mean±SD)** | 102.7 ± 7.0 | 102.4 ± 7.1 | 102.6 ± 7.0 | 0.04 | 99.4 ± 9.0 | 101.0 ± 6.5 | 100.5 ± 7.4 | 0.005 |
| **Latest creatinine^A^, mg/dL, (mean±SD)** | 1.7 ± 1.7 | 1.7 ± 1.6 | 1.7 ± 1.6 | 0.9 | 1.6 ± 1.6 | 1.4 ± 1.3 | 1.5 ± 1.4 | 0.005 |
| **Maximum creatinine^A^, mg/dL, (mean±SD)** | 1.7 ± 1.7 | 1.7 ± 1.6 | 1.7 ± 1.7 | 1.0 | 1.7 ± 1.6 | 1.5 ± 1.3 | 1.5 ± 1.4 | 0.009 |
| **Minimum creatinine^A^, mg/dL, (mean±SD)** | 1.6 ± 1.6 | 1.6 ± 1.6 | 1.6 ± 1.6 | 1.0 | 1.6 ± 1.6 | 1.4 ± 1.2 | 1.4 ± 1.4 | 0.005 |
| **Latest sodium^A^, mEq/L, (mean±SD)** | 138.7 ± 5.7 | 138.3 ± 5.8 | 138.5 ± 5.8 | < 0.001 | 139.8 ± 5.1 | 140.1 ± 5.0 | 140.0 ± 5.1 | 0.3 |
| **Maximum sodium^A^, mEq/L, (mean±SD)** | 139.7 ± 5.9 | 139.3 ± 5.9 | 139.5 ± 5.9 | 0.002 | 140.2 ± 5.2 | 140.5 ± 5.0 | 140.4 ± 5.1 | 0.1 |
| **Minimum sodium^A^, mEq/L, (mean±SD)** | 137.4 ± 5.9 | 137.0 ± 6.0 | 137.2 ± 5.9 | 0.002 | 138.9 ± 5.3 | 139.0 ± 5.2 | 138.9 ± 5.2 | 0.9 |
| **Latest potassium^A^, mEq/L, (mean±SD)** | 4.3 ± 0.7 | 4.3 ± 0.8 | 4.3 ± 0.7 | 0.8 | 4.2 ± 0.8 | 4.3 ± 0.7 | 4.2 ± 0.7 | 0.2 |
| **Maximum potassium^A^, mEq/L, (mean±SD)** | 4.5 ± 0.8 | 4.5 ± 0.9 | 4.5 ± 0.8 | 0.9 | 4.2 ± 0.8 | 4.3 ± 0.7 | 4.3 ± 0.7 | 0.1 |
| **Minimum potassium^A^, mEq/L, (mean±SD)** | 4.1 ± 0.7 | 4.0 ± 0.7 | 4.1 ± 0.7 | 0.2 | 4.2 ± 0.8 | 4.2 ± 0.7 | 4.2 ± 0.7 | 0.6 |
| **Latest INR^A^, (mean±SD)** | 1.6 ± 0.9 | 1.6 ± 0.9 | 1.6 ± 0.9 | 0.8 | 1.3 ± 0.4 | 1.4 ± 0.4 | 1.3 ± 0.4 | 0.09 |
| **Maximum INR^A^, (mean±SD)** | 1.7 ± 1.1 | 1.7 ± 1.0 | 1.7 ± 1.1 | 0.6 | 1.3 ± 0.4 | 1.4 ± 0.6 | 1.4 ± 0.5 | < 0.001 |
| **Minimum INR^A^, (mean±SD)** | 1.5 ± 0.9 | 1.5 ± 0.8 | 1.5 ± 0.8 | 1.0 | 1.3 ± 0.4 | 1.3 ± 0.3 | 1.3 ± 0.4 | 0.5 |
| **Latest PO2^A^, mm Hg, (mean±SD)** | 133.0 ± 72.1 | 128.4 ± 68.0 | 130.9 ± 70.3 | 0.006 | 95.6 ± 28.1 | 95.3 ± 27.9 | 95.4 ± 27.9 | 0.8 |
| **Maximum PO2^A^, mm Hg, (mean±SD)** | 215.4 ± 134.2 | 218.3 ± 135.1 | 216.7 ± 134.6 | 0.4 | 147.7 ± 78.1 | 182.8 ± 104.6 | 171.8 ± 98.4 | < 0.001 |
| **Minimum PO2^A^, mm Hg, (mean±SD)** | 115.3 ± 66.2 | 107.8 ± 58.2 | 111.9 ± 62.8 | < 0.001 | 77.9 ± 21.6 | 75.5 ± 19.6 | 76.2 ± 20.3 | 0.02 |
| **Latest PCO2^A^, mm Hg, (mean±SD)** | 41.3 ± 11.1 | 40.7 ± 10.1 | 41.0 ± 10.7 | 0.02 | 41.9 ± 10.4 | 40.5 ± 9.1 | 40.9 ± 9.5 | 0.006 |
| **Maximum PCO2^A^, mm Hg, (mean±SD)** | 47.0 ± 15.2 | 47.2 ± 14.2 | 47.1 ± 14.8 | 0.7 | 49.6 ± 14.9 | 49.1 ± 13.8 | 49.3 ± 14.2 | 0.5 |
| **Minimum PCO2^A^, mm Hg, (mean±SD)** | 38.3 ± 10.7 | 37.6 ± 9.9 | 38.0 ± 10.4 | 0.01 | 36.9 ± 9.4 | 35.0 ± 8.1 | 35.6 ± 8.6 | < 0.001 |
| **Net fluid balance^B^, mL, (mean±SD)** | 1685.2 ± 2748.4 | 2052.5 ± 2968.3 | 1846.8 ± 2852.9 | < 0.001 | 1941.2 ± 1948.5 | 2917.0 ± 2573.6 | 2613.8 ± 2438.6 | < 0.001 |
| **Urine output^B^, mL, (mean±SD)** | 1051.9 ± 1266.8 | 1011.0 ± 1361.0 | 1033.9 ± 1309.2 | 0.1 | 692.5 ± 856.3 | 889.9 ± 1140.3 | 828.6 ± 1063.9 | < 0.001 |

Abbreviation: ICU (intensive care unit), AKI (acute kidney injury), SBP (systolic blood pressure), DBP (diastolic blood pressure), MBP (mean blood pressure), SpO2 (oxygen saturation), MCH (mean corpuscular hemoglobin), MCHC (mean corpuscular hemoglobin concentration), MCV (mean corpuscular volume), RBC (red blood cell count), RDW (red cell distribution width), WBC (white blood cell count), BUN (blood urea nitrogen), INR (international normalised ratio), PO2 (partial pressure of oxygen), PCO2 (partial pressure of carbon dioxide)

^A^: Measurements within the Latest 48 hours prior to AKI onset

^B^: Measurements from ICU admission to AKI onset

**Supplementary Table S3.** Comparison of additional baseline characteristics of patients who actually received versus did not receive restrictive fluids within the subgroup predicted to benefit from restrictive fluids

|  | **Development cohort (MIMIC-IV)** | | | | **External validation cohort (SICdb)** | | | |
| --- | --- | --- | --- | --- | --- | --- | --- | --- |
|  | **Did not receive restrictive IV fluids**  **(N= 4,326)** | **Received restrictive IV fluids**  **(N= 798)** | **Total**  **(N= 5,124)** | **p value** | **Did not receive restrictive IV fluids**  **(N= 1255)** | **Received restrictive IV fluids**  **(N= 76)** | **Total**  **(N= 1331)** | **p value** |
| **The time (hour) between ICU admission and AKI onset, (mean±SD)** | 14.1 ± 10.0 | 16.4 ± 11.2 | 14.5 ± 10.2 | < 0.001 | 13.0 ± 9.7 | 17.2 ± 11.5 | 13.3 ± 9.9 | < 0.001 |
| **Latest heart rate^A^, (mean±SD)** | 88.6 ± 21.0 | 84.8 ± 20.6 | 88.0 ± 21.0 | < 0.001 | 86.8 ± 19.4 | 79.6 ± 17.8 | 86.4 ± 19.4 | 0.002 |
| **Maximum heart rate^A^, (mean±SD)** | 104.9 ± 23.6 | 100.1 ± 22.6 | 104.2 ± 23.5 | < 0.001 | 137.5 ± 42.4 | 124.6 ± 33.7 | 136.7 ± 42.0 | 0.009 |
| **Minimum heart rate^A^, (mean±SD)** | 76.7 ± 19.4 | 74.1 ± 18.2 | 76.2 ± 19.2 | < 0.001 | 61.9 ± 17.7 | 57.9 ± 14.4 | 61.7 ± 17.5 | 0.05 |
| **Latest SBP^A^, mmHg, (mean±SD)** | 111.8 ± 20.8 | 114.3 ± 21.6 | 112.2 ± 21.0 | 0.002 | 104.1 ± 20.5 | 113.0 ± 20.8 | 104.7 ± 20.6 | < 0.001 |
| **Maximum SBP^A^, mmHg, (mean±SD)** | 141.9 ± 25.2 | 143.1 ± 24.1 | 142.1 ± 25.0 | 0.2 | 217.2 ± 66.0 | 226.1 ± 63.5 | 217.7 ± 65.9 | 0.3 |
| **Minimum SBP^A^, mmHg, (mean±SD)** | 87.9 ± 18.5 | 90.8 ± 20.3 | 88.4 ± 18.8 | < 0.001 | 54.3 ± 22.0 | 62.7 ± 22.0 | 54.8 ± 22.1 | 0.001 |
| **Latest DBP^A^, mmHg, (mean±SD)** | 60.0 ± 14.1 | 61.0 ± 15.7 | 60.1 ± 14.4 | 0.06 | 53.5 ± 10.3 | 56.5 ± 10.8 | 53.7 ± 10.3 | 0.02 |
| **Maximum DBP^A^, mmHg, (mean±SD)** | 83.1 ± 21.3 | 85.1 ± 20.6 | 83.4 ± 21.2 | 0.01 | 130.5 ± 65.6 | 131.8 ± 62.3 | 130.5 ± 65.4 | 0.9 |
| **Minimum DBP^A^, mmHg, (mean±SD)** | 46.1 ± 12.1 | 47.3 ± 13.5 | 46.3 ± 12.3 | 0.01 | 24.6 ± 13.2 | 26.2 ± 12.5 | 24.7 ± 13.2 | 0.3 |
| **Latest MBP^A^, mmHg, (mean±SD)** | 75.0 ± 15.0 | 76.1 ± 15.5 | 75.2 ± 15.1 | 0.04 | 70.6 ± 12.5 | 75.6 ± 12.0 | 70.9 ± 12.6 | < 0.001 |
| **Maximum MBP^A^, mmHg, (mean±SD)** | 100.8 ± 26.8 | 102.5 ± 26.4 | 101.0 ± 26.7 | 0.09 | 176.3 ± 61.3 | 170.8 ± 51.8 | 176.0 ± 60.8 | 0.4 |
| **Minimum MBP^A^, mmHg, (mean±SD)** | 57.6 ± 14.7 | 60.2 ± 15.0 | 58.0 ± 14.8 | < 0.001 | 28.9 ± 18.2 | 28.9 ± 19.0 | 28.9 ± 18.2 | 1.0 |
| **Latest respiratory rate^A^, (mean±SD)** | 20.7 ± 5.8 | 20.7 ± 5.4 | 20.7 ± 5.8 | 0.8 | 15.8 ± 5.3 | 17.0 ± 6.5 | 15.9 ± 5.4 | 0.09 |
| **Maximum respiratory rate^A^, (mean±SD)** | 27.5 ± 7.0 | 27.4 ± 6.5 | 27.5 ± 6.9 | 0.8 | 30.3 ± 13.5 | 31.9 ± 13.6 | 30.4 ± 13.5 | 0.4 |
| **Minimum respiratory rate^A^, (mean±SD)** | 14.4 ± 4.7 | 14.4 ± 4.3 | 14.4 ± 4.7 | 0.9 | 8.3 ± 4.8 | 9.0 ± 4.4 | 8.3 ± 4.7 | 0.2 |
| **Latest temperature^A^, C, (mean±SD)** | 36.9 ± 0.8 | 36.9 ± 0.6 | 36.9 ± 0.8 | 0.2 | 36.7 ± 2.0 | 36.4 ± 2.6 | 36.7 ± 2.1 | 0.2 |
| **Maximum temperature^A^, C, (mean±SD)** | 37.3 ± 0.9 | 37.3 ± 0.7 | 37.3 ± 0.9 | 0.3 | 37.5 ± 1.1 | 37.6 ± 1.0 | 37.5 ± 1.1 | 0.7 |
| **Minimum temperature^A^, C, (mean±SD)** | 36.3 ± 1.0 | 36.4 ± 0.7 | 36.3 ± 0.9 | 0.1 | 31.8 ± 5.1 | 30.9 ± 5.9 | 31.7 ± 5.1 | 0.2 |
| **Latest SpO2^A^, %, (mean±SD)** | 96.8 ± 3.6 | 96.2 ± 3.4 | 96.7 ± 3.6 | < 0.001 | 96.2 ± 4.0 | 95.7 ± 2.8 | 96.2 ± 4.0 | 0.3 |
| **Maximum SpO2^A^, %, (mean±SD)** | 99.2 ± 2.0 | 99.1 ± 1.7 | 99.2 ± 1.9 | 0.03 | 99.7 ± 1.2 | 99.5 ± 1.1 | 99.7 ± 1.2 | 0.1 |
| **Minimum SpO2^A^, %, (mean±SD)** | 92.3 ± 6.8 | 91.9 ± 5.7 | 92.2 ± 6.7 | 0.2 | 80.6 ± 11.9 | 81.6 ± 9.8 | 80.6 ± 11.7 | 0.5 |
| **Weight^A^, Kg, (mean±SD)** | 85.4 ± 26.3 | 84.2 ± 22.8 | 85.2 ± 25.8 | 0.2 | 79.7 ± 22.6 | 83.3 ± 23.9 | 79.9 ± 22.6 | 0.2 |
| **Latest hematocrit^A^, %, (mean±SD)** | 32.0 ± 6.2 | 31.9 ± 6.7 | 32.0 ± 6.3 | 0.7 | 30.3 ± 5.8 | 31.2 ± 6.2 | 30.4 ± 5.8 | 0.2 |
| **Maximum hematocrit^A^, %, (mean±SD)** | 34.1 ± 6.6 | 33.9 ± 6.7 | 34.1 ± 6.3 | 0.4 | 32.4 ± 6.3 | 34.1 ± 6.0 | 32.5 ± 6.2 | 0.03 |
| **Minimum hematocrit^A^, %, (mean±SD)** | 30.7 ± 6.8 | 30.9 ± 7.1 | 30.7 ± 6.8 | 0.6 | 29.6 ± 6.1 | 30.7 ± 6.4 | 29.7 ± 6.1 | 0.2 |
| **Latest hemoglobin^A^, g/dL, (mean±SD)** | 10.6 ± 2.1 | 10.4 ± 2.2 | 10.6 ± 2.1 | 0.06 | 10.3 ± 2.0 | 10.5 ± 2.1 | 10.3 ± 2.0 | 0.4 |
| **Maximum hemoglobin^A^, g/dL, (mean±SD)** | 11.2 ± 2.1 | 11.0 ± 2.3 | 11.2 ± 2.1 | 0.03 | 11.0 ± 2.2 | 11.5 ± 2.2 | 11.1 ± 2.2 | 0.06 |
| **Minimum hemoglobin^A^, g/dL, (mean±SD)** | 10.1 ± 2.2 | 10.1 ± 2.4 | 10.1 ± 2.3 | 0.4 | 10.1 ± 2.1 | 10.4 ± 2.2 | 10.1 ± 2.1 | 0.2 |
| **Latest MCH^A^, pg, (mean±SD)** | 30.2 ± 2.7 | 29.9 ± 2.8 | 30.1 ± 2.7 | 0.02 | 30.3 ± 2.4 | 30.0 ± 2.8 | 30.2 ± 2.4 | 0.4 |
| **Maximum MCH^A^, pg, (mean±SD)** | 30.4 ± 2.8 | 30.2 ± 2.9 | 30.4 ± 2.8 | 0.02 | 30.4 ± 2.4 | 30.3 ± 2.9 | 30.4 ± 2.5 | 0.6 |
| **Minimum MCH^A^, pg, (mean±SD)** | 29.9 ± 2.7 | 29.7 ± 2.9 | 29.9 ± 2.8 | 0.02 | 30.1 ± 2.4 | 29.8 ± 2.9 | 30.1 ± 2.5 | 0.4 |
| **Latest MCHC^A^, g/L, (mean±SD)** | 33.0 ± 1.8 | 32.6 ± 1.8 | 32.9 ± 1.8 | < 0.001 | 34.0 ± 1.5 | 33.8 ± 1.4 | 34.0 ± 1.5 | 0.2 |
| **Maximum MCHC^A^, g/L, (mean±SD)** | 33.3 ± 1.9 | 32.8 ± 1.8 | 33.2 ± 1.9 | < 0.001 | 34.3 ± 1.5 | 34.1 ± 1.6 | 34.3 ± 1.5 | 0.4 |
| **Minimum MCHC^A^, g/L, (mean±SD)** | 32.6 ± 1.7 | 32.2 ± 1.7 | 32.5 ± 1.7 | < 0.001 | 33.8 ± 1.5 | 33.5 ± 1.4 | 33.8 ± 1.5 | 0.1 |
| **Latest MCV^A^, fL, (mean±SD)** | 91.5 ± 7.3 | 91.9 ± 7.3 | 91.6 ± 7.3 | 0.2 | 88.9 ± 6.2 | 88.9 ± 7.8 | 88.9 ± 6.3 | 1.0 |
| **Maximum MCV^A^, fL, (mean±SD)** | 92.5 ± 7.4 | 92.8 ± 7.6 | 92.6 ± 7.4 | 0.3 | 89.5 ± 6.3 | 89.5 ± 7.9 | 89.5 ± 6.4 | 1.0 |
| **Minimum MCV^A^, fL, (mean±SD)** | 91.0 ± 7.3 | 91.3 ± 7.3 | 91.0 ± 7.3 | 0.2 | 88.4 ± 6.2 | 88.3 ± 7.9 | 88.4 ± 6.3 | 1.0 |
| **Latest platelet^A^, K/uL, (mean±SD)** | 199.9 ± 115.9 | 200.7 ± 103.4 | 200.1 ± 114.0 | 0.9 | 197.5 ± 101.1 | 236.7 ± 105.5 | 199.7 ± 101.7 | 0.002 |
| **Maximum platelet^A^, K/uL, (mean±SD)** | 220.0 ± 122.1 | 217.6 ± 106.3 | 219.7 ± 119.8 | 0.6 | 218.4 ± 107.2 | 266.3 ± 119.8 | 221.1 ± 108.5 | < 0.001 |
| **Minimum platelet^A^, K/uL, (mean±SD)** | 189.8 ± 115.1 | 191.6 ± 100.5 | 190.1 ± 112.9 | 0.7 | 191.2 ± 100.3 | 230.1 ± 107.4 | 193.5 ± 101.1 | 0.002 |
| **Latest RBC^A^, Count m/uL, (mean±SD)** | 3.5 ± 0.7 | 3.5 ± 0.8 | 3.5 ± 0.7 | 0.4 | 3.4 ± 0.6 | 3.5 ± 0.7 | 3.4 ± 0.6 | 0.2 |
| **Maximum RBC^A^ Count, m/uL, (mean±SD)** | 3.7 ± 0.7 | 3.7 ± 0.8 | 3.7 ± 0.7 | 0.3 | 3.7 ± 0.7 | 3.9 ± 0.7 | 3.7 ± 0.7 | 0.02 |
| **Minimum RBC^A^ Count, m/uL, (mean±SD)** | 3.4 ± 0.8 | 3.4 ± 0.8 | 3.4 ± 0.8 | 0.9 | 3.3 ± 0.7 | 3.5 ± 0.7 | 3.3 ± 0.7 | 0.1 |
| **Latest WBC^A^, K/uL, (mean±SD)** | 13.8 ± 10.0 | 13.2 ± 8.0 | 13.7 ± 9.7 | 0.1 | 12.6 ± 7.3 | 12.0 ± 6.2 | 12.5 ± 7.2 | 0.5 |
| **Maximum WBC^A^, K/uL, (mean±SD)** | 15.3 ± 10.5 | 14.6 ± 8.4 | 15.2 ± 10.2 | 0.07 | 13.9 ± 7.6 | 13.6 ± 6.3 | 13.9 ± 7.5 | 0.8 |
| **Minimum WBC^A^, K/uL, (mean±SD)** | 12.2 ± 9.5 | 11.6 ± 7.4 | 12.1 ± 9.2 | 0.2 | 11.1 ± 6.6 | 10.8 ± 5.9 | 11.1 ± 6.6 | 0.7 |
| **Latest anion gap^A^, mEq/L, (mean±SD)** | 15.1 ± 4.6 | 14.2 ± 4.4 | 15.0 ± 4.6 | < 0.001 | 13.9 ± 4.3 | 13.6 ± 3.8 | 13.9 ± 4.3 | 0.7 |
| **Maximum anion gap^A^, mEq/L, (mean±SD)** | 16.5 ± 4.9 | 15.8 ± 4.7 | 16.4 ± 4.8 | < 0.001 | 17.0 ± 4.4 | 15.9 ± 3.3 | 16.9 ± 4.4 | 0.2 |
| **Minimum anion gap^A^, mEq/L, (mean±SD)** | 14.1 ± 4.5 | 13.2 ± 4.4 | 14.0 ± 4.5 | < 0.001 | 12.1 ± 5.0 | 11.9 ± 4.8 | 12.1 ± 4.9 | 0.8 |
| **Latest bicarbonate^A^, mEq/L, (mean±SD)** | 22.0 ± 5.0 | 22.9 ± 5.4 | 22.1 ± 5.1 | < 0.001 | 23.9 ± 4.4 | 26.0 ± 4.6 | 24.0 ± 4.4 | < 0.001 |
| **Maximum bicarbonate^A^, mEq/L, (mean±SD)** | 23.1 ± 5.1 | 24.0 ± 5.4 | 23.2 ± 5.1 | < 0.001 | 25.7 ± 4.7 | 27.4 ± 4.6 | 25.8 ± 4.7 | 0.004 |
| **Minimum bicarbonate^A^, mEq/L, (mean±SD)** | 21.0 ± 5.1 | 21.7 ± 5.6 | 21.1 ± 5.2 | < 0.001 | 20.8 ± 4.4 | 22.6 ± 4.2 | 20.9 ± 4.4 | < 0.001 |
| **Latest BUN^A^, mg/dL, (mean±SD)** | 30.6 ± 23.8 | 34.6 ± 25.6 | 31.3 ± 24.1 | < 0.001 | 26.1 ± 20.8 | 23.2 ± 21.5 | 25.9 ± 20.8 | 0.3 |
| **Maximum BUN^A^, mg/dL, (mean±SD)** | 32.0 ± 24.1 | 36.1 ± 26.1 | 32.6 ± 24.5 | < 0.001 | 27.0 ± 21.2 | 24.6 ± 21.7 | 26.9 ± 21.3 | 0.4 |
| **Minimum BUN^A^, mg/dL, (mean±SD)** | 29.2 ± 23.6 | 32.6 ± 25.2 | 29.8 ± 23.9 | < 0.001 | 25.1 ± 20.9 | 22.2 ± 21.7 | 24.9 ± 20.9 | 0.3 |
| **Latest calcium^A^, mg/dL, (mean±SD)** | 8.1 ± 0.9 | 8.5 ± 0.9 | 8.9 ± 0.9 | < 0.001 | 8.4 ± 0.6 | 8.5 ± 0.6 | 8.4 ± 0.6 | 0.1 |
| **Maximum calcium^A^, mg/dL, (mean±SD)** | 8.4 ± 1.0 | 8.7 ± 0.9 | 8.4 ± 0.9 | < 0.001 | 8.6 ± 0.7 | 8.8 ± 0.6 | 8.6 ± 0.7 | 0.009 |
| **Minimum calcium^A^, mg/dL, (mean±SD)** | 7.9 ± 1.0 | 8.3 ± 0.8 | 8.0 ± 0.9 | < 0.001 | 8.4 ± 0.6 | 8.5 ± 0.6 | 8.4 ± 0.6 | 0.2 |
| **Latest chloride^A^, mEq/L, (mean±SD)** | 104.4 ± 7.1 | 102.4 ± 6.5 | 104.1 ± 7.0 | < 0.001 | 101.5 ± 6.2 | 100.9 ± 5.1 | 101.5 ± 6.2 | 0.6 |
| **Maximum chloride^A^, mEq/L, (mean±SD)** | 105.5 ± 7.4 | 103.6 ± 6.7 | 105.2 ± 7.3 | < 0.001 | 101.6 ± 6.3 | 101.0 ± 5.0 | 101.6 ± 6.2 | 0.6 |
| **Minimum chloride^A^, mEq/L, (mean±SD)** | 102.7 ± 7.2 | 100.7 ± 6.6 | 102.4 ± 7.1 | < 0.001 | 101.0 ± 6.5 | 100.4 ± 5.1 | 101.0 ± 6.5 | 0.6 |
| **Latest creatinine^A^, mg/dL, (mean±SD)** | 1.6 ± 1.5 | 2.0 ± 2.0 | 1.7 ± 1.6 | < 0.001 | 1.4 ± 1.2 | 1.3 ± 1.3 | 1.4 ± 1.3 | 0.3 |
| **Maximum creatinine^A^, mg/dL, (mean±SD)** | 1.7 ± 1.5 | 2.0 ± 2.0 | 1.7 ± 1.6 | < 0.001 | 1.5 ± 1.3 | 1.4 ± 1.3 | 1.5 ± 1.3 | 0.4 |
| **Minimum creatinine^A^, mg/dL, (mean±SD)** | 1.5 ± 1.5 | 1.8 ± 1.9 | 1.6 ± 1.6 | < 0.001 | 1.4 ± 1.2 | 1.2 ± 1.3 | 1.4 ± 1.2 | 0.3 |
| **Latest sodium^A^, mEq/L, (mean±SD)** | 138.4 ± 5.9 | 137.8 ± 5.5 | 138.3 ± 5.8 | 0.01 | 140.0 ± 5.0 | 140.7 ± 5.0 | 140.1 ± 5.0 | 0.3 |
| **Maximum sodium^A^, mEq/L, (mean±SD)** | 139.4 ± 6.0 | 138.8 ± 5.5 | 139.3 ± 5.9 | 0.02 | 140.5 ± 5.0 | 141.0 ± 4.9 | 140.5 ± 5.0 | 0.4 |
| **Minimum sodium^A^, mEq/L, (mean±SD)** | 137.1 ± 6.1 | 136.4 ± 5.6 | 137.0 ± 6.0 | < 0.001 | 138.9 ± 5.2 | 139.3 ± 4.9 | 139.0 ± 5.2 | 0.6 |
| **Latest potassium^A^, mEq/L, (mean±SD)** | 4.3 ± 0.7 | 4.4 ± 0.8 | 4.3 ± 0.8 | < 0.001 | 4.3 ± 0.7 | 4.2 ± 0.7 | 4.3 ± 0.7 | 0.7 |
| **Maximum potassium^A^, mEq/L, (mean±SD)** | 4.5 ± 0.8 | 4.7 ± 0.9 | 4.5 ± 0.9 | < 0.001 | 4.3 ± 0.7 | 4.3 ± 0.7 | 4.3 ± 0.7 | 0.8 |
| **Minimum potassium^A^, mEq/L, (mean±SD)** | 4.0 ± 0.7 | 4.1 ± 0.8 | 4.0 ± 0.7 | 0.003 | 4.2 ± 0.7 | 4.2 ± 0.6 | 4.2 ± 0.7 | 0.8 |
| **Latest INR^A^, (mean±SD)** | 1.6 ± 0.8 | 1.6 ± 1.0 | 1.6 ± 0.9 | 0.2 | 1.4 ± 0.4 | 1.2 ± 0.1 | 1.4 ± 0.4 | 0.02 |
| **Maximum INR^A^, (mean±SD)** | 1.7 ± 1.0 | 1.7 ± 1.1 | 1.7 ± 1.0 | 0.8 | 1.4 ± 0.6 | 1.3 ± 0.1 | 1.4 ± 0.6 | 0.02 |
| **Minimum INR^A^, (mean±SD)** | 1.5 ± 0.8 | 1.5 ± 0.9 | 1.5 ± 0.8 | 0.1 | 1.3 ± 0.4 | 1.2 ± 0.1 | 1.3 ± 0.3 | 0.05 |
| **Latest PO2^A^, mm Hg, (mean±SD)** | 129.5 ± 68.6 | 120.6 ± 63.6 | 128.4 ± 68.0 | 0.02 | 95.5 ± 27.4 | 91.8 ± 35.2 | 95.3 ± 27.9 | 0.3 |
| **Maximum PO2^A^, mm Hg, (mean±SD)** | 220.7 ± 135.2 | 201.3 ± 133.3 | 218.3 ± 135.1 | 0.008 | 183.7 ± 105.0 | 168.3 ± 97.4 | 182.8 ± 104.6 | 0.2 |
| **Minimum PO2^A^, mm Hg, (mean±SD)** | 108.5 ± 58.8 | 102.7 ± 53.5 | 107.8 ± 58.2 | 0.06 | 75.7 ± 19.9 | 71.5 ± 14.0 | 75.5 ± 19.6 | 0.09 |
| **Latest PCO2^A^, mm Hg, (mean±SD)** | 40.4 ± 9.8 | 42.6 ± 11.9 | 40.7 ± 10.1 | < 0.001 | 40.4 ± 9.0 | 41.8 ± 10.4 | 40.5 ± 9.1 | 0.2 |
| **Maximum PCO2^A^, mm Hg, (mean±SD)** | 47.0 ± 14.1 | 48.3 ± 15.1 | 47.2 ± 14.2 | 0.1 | 49.0 ± 13.9 | 50.2 ± 13.0 | 49.1 ± 13.8 | 0.5 |
| **Minimum PCO2^A^, mm Hg, (mean±SD)** | 37.4 ± 9.6 | 39.7 ± 11.7 | 37.6 ± 9.9 | < 0.001 | 35.0 ± 8.1 | 35.9 ± 8.1 | 35.0 ± 8.1 | 0.4 |
| **Net fluid balance^B^, mL, (mean±SD)** | 2470.7 ± 2940.1 | -214.9 ± 1905.0 | 2052.5 ± 2968.3 | < 0.001 | 3007.0 ± 2571.5 | 1430.6 ± 2125.5 | 2917.0 ± 2573.6 | < 0.001 |
| **Urine output^B^, mL, (mean±SD)** | 979.3 ± 1358.3 | 1182.9 ± 1364.0 | 1011.0 ± 1361.0 | < 0.001 | 870.6 ± 1143.9 | 1209.5 ± 1034.2 | 889.9 ± 1140.3 | 0.01 |

Abbreviation: ICU (intensive care unit), AKI (acute kidney injury), SBP (systolic blood pressure), DBP (diastolic blood pressure), MBP (mean blood pressure), SpO2 (oxygen saturation), MCH (mean corpuscular hemoglobin), MCHC (mean corpuscular hemoglobin concentration), MCV (mean corpuscular volume), RBC (red blood cell count), RDW (red cell distribution width), WBC (white blood cell count), BUN (blood urea nitrogen), INR (international normalised ratio), PO2 (partial pressure of oxygen), PCO2 (partial pressure of carbon dioxide)

^A^: Measurements within the Latest 48 hours prior to AKI onset

^B^: Measurements from ICU admission to AKI onset

**Supplementary Table S4.** Sensitivity analysis showing impact of restrictive fluid strategy among patients with congestive heart failure who were predicted to benefit from restrictive fluids

|  | Development Cohort (MIMIC-IV) | | | External Validation Cohort (SICdb) | | |
| --- | --- | --- | --- | --- | --- | --- |
|  | Did not receive restrictive IV fluids  (N = 2598) | Received restrictive IV fluids  (N = 668) | p value | Did not receive restrictive IV Fluids | Received restrictive IV Fluids | p value |
| Early AKI Reversal | 850 (32.7%) | 386 (57.8%) | < 0.001 | NA | NA | NA |
| Sustained AKI Reversal | 352 (13.5%) | 157 (23.5%) | < 0.001 | NA | NA | NA |
| MAKE30 | 843 (32.4%) | 176 (26.3%) | 0.003 | NA | NA | NA |

* History of congestive heart failure was not available in SICdb.

**Supplementary Table S5.** Sensitivity analysis showing impact of restrictive fluid strategy among patients with chronic kidney disease who were predicted to benefit from restrictive fluids

|  | Development Cohort (MIMIC-IV) | | | External Validation Cohort (SICdb) | | |
| --- | --- | --- | --- | --- | --- | --- |
|  | Did not receive restrictive IV fluids  (N = 807) | Received restrictive IV fluids  (N = 202) | p value | Did not receive restrictive IV fluids  (N = 189) | Received restrictive IV fluids  (N = 14) | p value |
| Early AKI Reversal | 222 (27.5%) | 101 (50.0%) | < 0.001 | 22 (19.0%) | 4 (33.3%) | 0.4 |
| Sustained AKI Reversal | 98 (12.1%) | 44 (21.8%) | < 0.001 | 12 (10.3%) | 3 (25.0%) | 0.3 |
| MAKE30 | 262 (32.5%) | 43 (21.3%) | 0.003 | 56 (48.3%) | 3 (25.0%) | 0.2 |

**Supplementary Table S6.** Sensitivity analysis showing impact of restrictive fluid strategy among patients who were predicted to benefit from restrictive fluids when admission creatinine was treated as baseline creatinine for all patients

|  | Development Cohort (MIMIC-IV) | | | External Validation Cohort (SICdb) | | |
| --- | --- | --- | --- | --- | --- | --- |
|  | Did not receive restrictive IV fluids  (N = 4,268) | Received restrictive IV fluids  (N = 807) | p value | Did not receive restrictive IV fluids  (N = 1,200) | Received restrictive IV fluids  (N = 71) | p value |
| Early AKI Reversal | 1589 (37.2%) | 431 (53.4%) | < 0.001 | 432 (36.0%) | 40 (56.3%) | < 0.001 |
| Sustained AKI Reversal | 751 (17.6%) | 189 (23.4%) | < 0.001 | 238 (19.8%) | 24 (33.8%) | 0.007 |
| MAKE30 | 1127 (26.4%) | 172 (21.3%) | 0.003 | 365 (30.4%) | 12 (16.9%) | 0.02 |
